# Disorder-specific and shared genetic architecture underlying schizophrenia and bipolar disorder

**DOI:** 10.64898/2026.07.07.26357436

**Authors:** Upasana Bhattacharyya, Jibin John, Michael Preuss, Max Lam, Todd Lencz

## Abstract

Schizophrenia (SCZ) and bipolar disorder (BIP) share substantial genetic liability, yet exhibit heterogeneity in cognition, illness course and cardiometabolic health. Conventional disorder-level genome-wide association studies (GWASs) aggregate genetic liability obscuring this heterogeneity.

We applied zero-phase component analysis whitening and common factor GWAS to SCZ and BIP summary statistics, deriving a shared component (PSY_shared_) and two mutually orthogonal, disorder-predominant components (SCZ_∼BIP_ and BIP_∼SCZ_). We then examined whether the three profiles differed in locus architecture, pathway enrichment, developmental expression and intermediate-phenotype associations.

The decomposition identified 261 loci, of which 201 (77%) were component-specific and 16 loci were not reported in any previous larger SCZ and BIP GWASs or multivariate psychosis GWASs. PSY_shared_ was enriched for synaptic architecture, nuclear organisation and protein trafficking, with developmental signals spanning early-and late-mid-prenatal stages, late infancy and young adulthood. SCZ_∼BIP_ implicated cortical neuronal differentiation, DNA/RNA regulation and protein turnover, with developmental associations in late infancy and young adulthood. BIP_∼SCZ_ showed synaptic release and transport signals, alongside exploratory associations with gene/protein regulation and mitochondrial translation (nominal P < 0.001), with significant developmental enrichment in late infancy. Correlations of SCZ and BIP with intermediate traits were asymmetric, whereas SCZ∼_BIP_and BIP∼_SCZ_diverged in opposite directions across cognitive performance, educational attainment, metabolic syndrome, and systemic inflammatory markers. In contrast, both orthogonal components correlated positively with risk taking, with a larger estimate for BIP_∼SCZ_; PSY_shared_ showed the largest estimate among the derived components. Steiger tests showed that the components differed significantly across all six phenotypes. Component contrasts were 1.63-2.07-fold greater than corresponding SCZ-BIP contrasts with the increase reaching significance for cognitive performance and metabolic syndrome.

These findings support a multidimensional representation of SCZ-BIP liability and provide a framework for testing whether relative component burdens account for variation in cognition, cardiometabolic and inflammatory burden, illness course, and treatment response beyond diagnosis.

## Introduction

Schizophrenia (SCZ) and bipolar disorder (BIP) are severe psychiatric disorders^1,2^ that usually begin in late adolescence or early adulthood^3,4^ and are highly heritable^5,6^. These two disorders are strongly genetically correlated (rg ≈ 0.70)^7^, reflecting extensive shared biology^8,9^, and have considerable phenotypic similarities^10,11^, yet show marked heterogeneity in clinical outcomes and other phenotypic features both within and across these diagnoses^12,13^. This paradoxical relationship suggests that each disorder may reflect shared liability together with relatively independent SCZ-or BIP-specific liability, and that their relative balance may contribute to diagnostic presentation and within-diagnosis heterogeneity.

Genome-wide association studies (GWAS) have identified growing numbers of common-variant loci^14,15^, including 370 for schizophrenia^16^ and 298 for bipolar disorder in multi-ancestry analyses^14^. However, conventional disorder-level GWASs identify aggregated common-variant liability for each diagnostic category. This is effective for locus discovery, but it averages genetic liability across potentially different routes to illness. A recently published review proposes that risk for complex diseases, including schizophrenia, is distributed across functionally distinguishable processes and may be better represented by a liability profile across multiple biological pathways rather than a single genome-wide score^17^. For example, genomic studies of Type 2 diabetes illustrate how advanced statistical approaches can identify dissociable mechanistic dimensions with different downstream associations^18,19^. In the context of schizophrenia and bipolar disorder, this insight motivates construction of a multidimensional map that extracts transdiagnostic liability while also resolving diagnosis-specific profiles.

Existing multivariate approaches, including Genomic SEM^20^, mtCOJO^21^ and CC-GWAS^22^, have been utilized to address broader questions concerning shared genetic architecture, disorder-conditioned associations and genetic differences between diagnostic groups; however, to date, no studies have been specifically designed to map biological heterogeneity against the relative contributions of genetic liability shared across the disorders and genetic liability predominantly aligned with SCZ or BIP.

Building on this premise, the present study aimed to derive two mutually orthogonal profiles (SCZ_∼BIP_ and BIP_∼SCZ_) that retained close alignment with the original diagnostic GWASs, together with a separately estimated shared-liability component (PSY_shared_). The purpose was to determine whether this decomposition could resolve biologically and phenotypically informative dimensions of liability, which in turn can help to clarify the heterogeneity observed within and between schizophrenia and bipolar disorder^23^. This approach may implicate loci and gene set pathways not recoverable from either input GWAS alone. In addition to comprehensive downstream characterization of summary statistics derived from the distinct liability components, we examined the relationship of the decomposed GWAS dimensions with six intermediate phenotypes spanning domains implicated in one or both disorders. Specifically, we examined GWAS data indexing cognitive processes (cognitive task performance and educational attainment^24,25^), behavioral disinhibition (risk taking^26,27^), cardiometabolic liability (metabolic syndrome^28–30^), and systemic inflammation^31,32^, indexed here by C-reactive protein^33,34,35^ and neutrophil percentage^36,37^. If the decomposition approach more accurately captures biological risk factors, then the derived components should provide more clearly resolved intermediate-phenotype relationships relative to the parent disorder GWASs.

We used Zero-phase Component Analysis (ZCA) to derive SCZ_∼BIP_ and BIP_∼SCZ_ from the LDSC genetic-covariance matrix, and common-factor GWAS implemented within Genomic SEM to estimate Psy_shared_ separately. ZCA was selected because it is invariant to trait order and, among whitening solutions, minimizes displacement from the input variables^38^. ZCA has been used in multi-trait GWAS methods to decorrelate vectors of z scores^39,40^, but, to our knowledge, has not been used to construct input-aligned orthogonal GWAS profiles from an LDSC genetic-covariance matrix. Therefore, in addition, we used bidirectional mtCOJO^21^ as a sensitivity benchmark. We then evaluated alignment with the source disorders and independent psychiatric endpoints, genomic loci and functional annotations, developmental expression, and relationships with the six intermediate phenotypes.

## Methods

### Overview of methodological framework

We used European-ancestry GWASs of schizophrenia (PGC3; 53,386 cases and 77,258 controls)^15^ and bipolar disorder (59,287 cases and 781,022 controls)^14^ to derive two orthogonal components and one shared component. We applied ZCA whitening to derive the input-aligned orthogonal SCZ and BIP components and used Genomic SEM^20^ to derive the SCZ-BIP common component, PSY (Figure 1). As noted above, we also used bidirectional mtCOJO ^21^ as an established benchmark for every analysis performed with a ZCA-derived orthogonal component. The Supplementary Methods describe component derivation, quality control, and sensitivity analyses. We functionally characterized each component through genomic risk-locus definition and candidate-gene mapping using the FUMA pipeline^41^; identification of component loci that were not genome-wide significant in the input GWASs using BEDTools^42^, competitive gene-set and BrainSpan analyses using MAGMA^43^ and synapse-specific enrichment using SynGO^44^. Component loci were compared with five published schizophrenia PGC GWASs^15,16,45–47^, three bipolar-disorder PGC GWASs^14,48,49^, and a shared SCZ-BIP factor from two recent Genomic SEM studies^8,9^.

**Figure 1.**
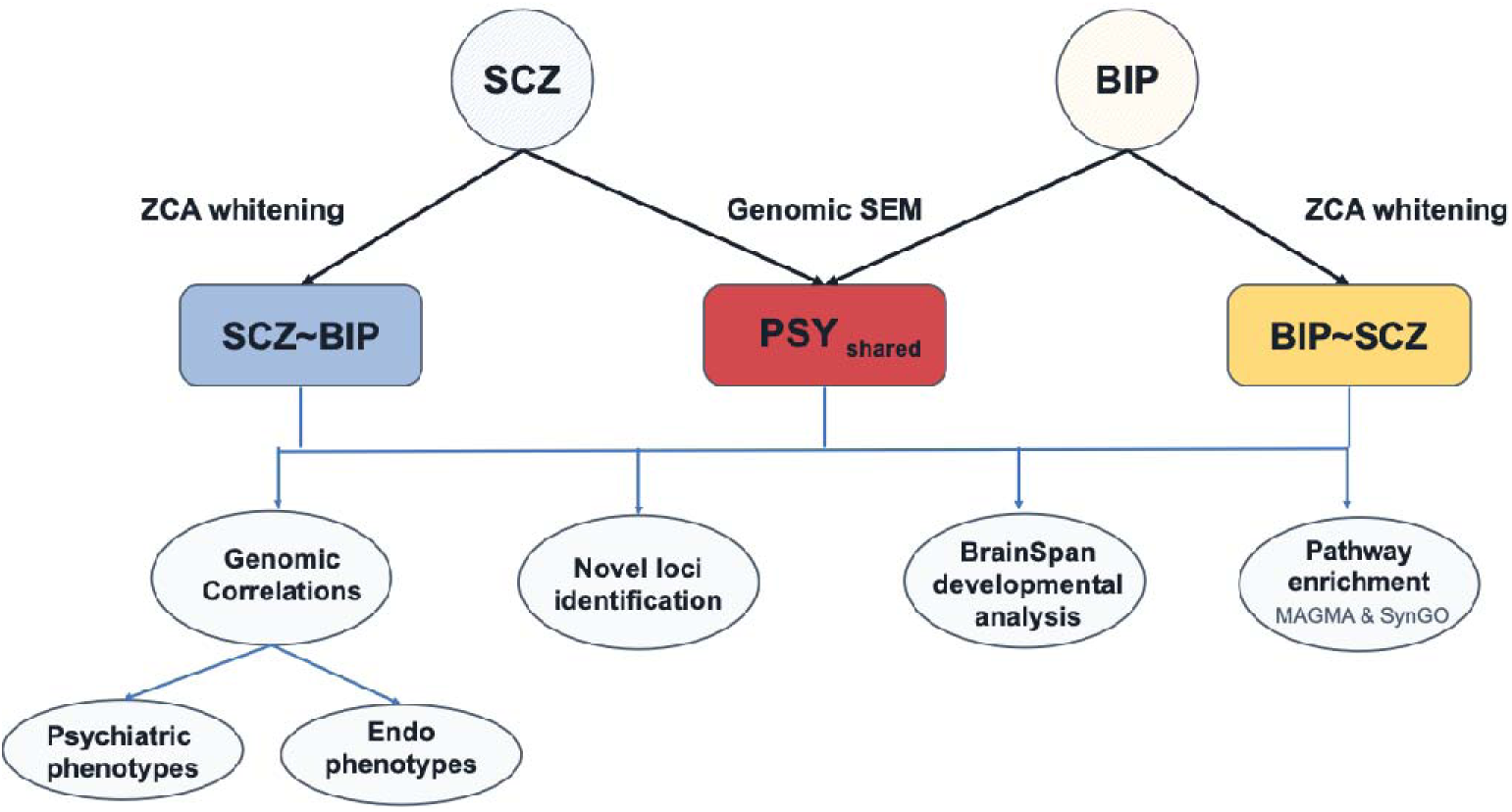
Study design: decomposition of schizophrenia and bipolar disorder GWASs into parent-aligned orthogonal and SCZ-BIP shared components and downstream characterization. Summary-statistic GWASs of SCZ and BIP were decomposed using ZCA whitening to derive SCZ_∼BIP_ and BIP_∼SCZ_ and Genomic SEM to derive Psy_shared_. Components were characterized by risk-locus and candidate-gene mapping, comparison with input-GWAS loci, MAGMA pathway and BrainSpan enrichment, SynGO analysis, and LDSC genetic correlations with psychiatric, cognitive, cardiometabolic, and inflammatory phenotypes. Covariance-aware pairwise tests and Wald comparisons evaluated component and SCZ-BIP contrasts.

We estimated inter-component genetic correlations and correlations with FinnGen R13 schizophrenia (7,312 cases; 484,297 controls), bipolar disorder (9,221 cases; 432,672 controls), schizoaffective disorder (3,168 cases; 484,297 controls), and other psychosis (9,216 cases; 484,297 controls) endpoints^50^, and with the schizophrenia-bipolar factor from the 14-disorder Genomic SEM model^9^. The six intermediate phenotypes GWASs comprised cognitive-task performance (N = 373,617)^51^, educational attainment (N = 766,345)^52^, risk taking (104,809 cases and 301,022 controls)^53^, metabolic syndrome (N_eff_ = 1,384,348)^54^, C-reactive protein (N = 575,531)^33^, and neutrophil percentage (N = 394,642)^55^. We used LDSC^56^ to estimate their genetic correlations with each component.

## Results

Decomposition generated SCZ_∼BIP_, BIP_∼SCZ_, and PSY_shared_, which differed in locus architecture, functional and developmental enrichment, and external genetic correlations, as detailed in the sections below. Bidirectional mtCOJO provided a sensitivity benchmark; its corresponding profiles (SCZ|BIP and BIP|SCZ) were highly correlated with the ZCA profiles (SCZ_∼BIP_ with SCZ|BIP, r_g_ = 0.994; BIP_∼SCZ_ with BIP|SCZ, r_g_ = 0.996) and showed concordant downstream patterns (Supplementary Results). ZCA was chosen as primary output because its components were empirically uncorrelated (r_g_ = −0.025), whereas the mtCOJO components retained modest negative correlation (r_g_ = −0.174). Both approaches remained aligned with the corresponding input GWASs and correlated positively with PSY_shared_ (Supplementary Fig. 1). Results are detailed below for the ZCA-derived components; as noted in the Supplementary text, results for mtCOJO-derived components were generally similar, although statistically weaker.

### SNP heritability

SNP heritability of the decomposed elements was 0.1377 (SE = 0.0059) for SCZ_∼BIP_, 0.0539 (SE = 0.0027) for BIP_∼SCZ_, and 0.1865 (SE = 0.0065) for PSY_shared_. For comparison, SNP heritability of the original, diagnosis-based GWAS was 0.2123 (SE = 0.0075) for SCZ and 0.0902 (SE = 0.0035) for BIP.

### Genomic risk loci and cross-component overlap

SCZ_∼BIP_ mapped to 98 independent genomewide significant risk loci, BIP_∼SCZ_ to 29, and PSY_shared_ to 197; the input SCZ and BIP GWASs were mapped to 175 and 74 loci, respectively (Figure 2a; Supplementary Table 1). The lower BIP_∼SCZ_ yield is consistent with the lower SNP heritability and locus yield of its input GWAS. Merging loci at ≥1-bp overlap produced 261 consensus loci (Figure 2b; Supplementary Table 1). Of these, 139 were unique to PSY_shared_, 46 to SCZ_∼BIP_, and 16 to BIP_∼SCZ_; thus, a total of 201 loci (77%) were component-specific. Forty-seven genomewide significant loci were shared by SCZ_∼BIP_ and PSY_shared_, 9 were shared by BIP_∼SCZ_ and PSY_shared_, 2 were shared by the two disease-specific components but not PSY_shared_, and 2 loci were significant in all three components (Figure 2b).

**Figure 2.**
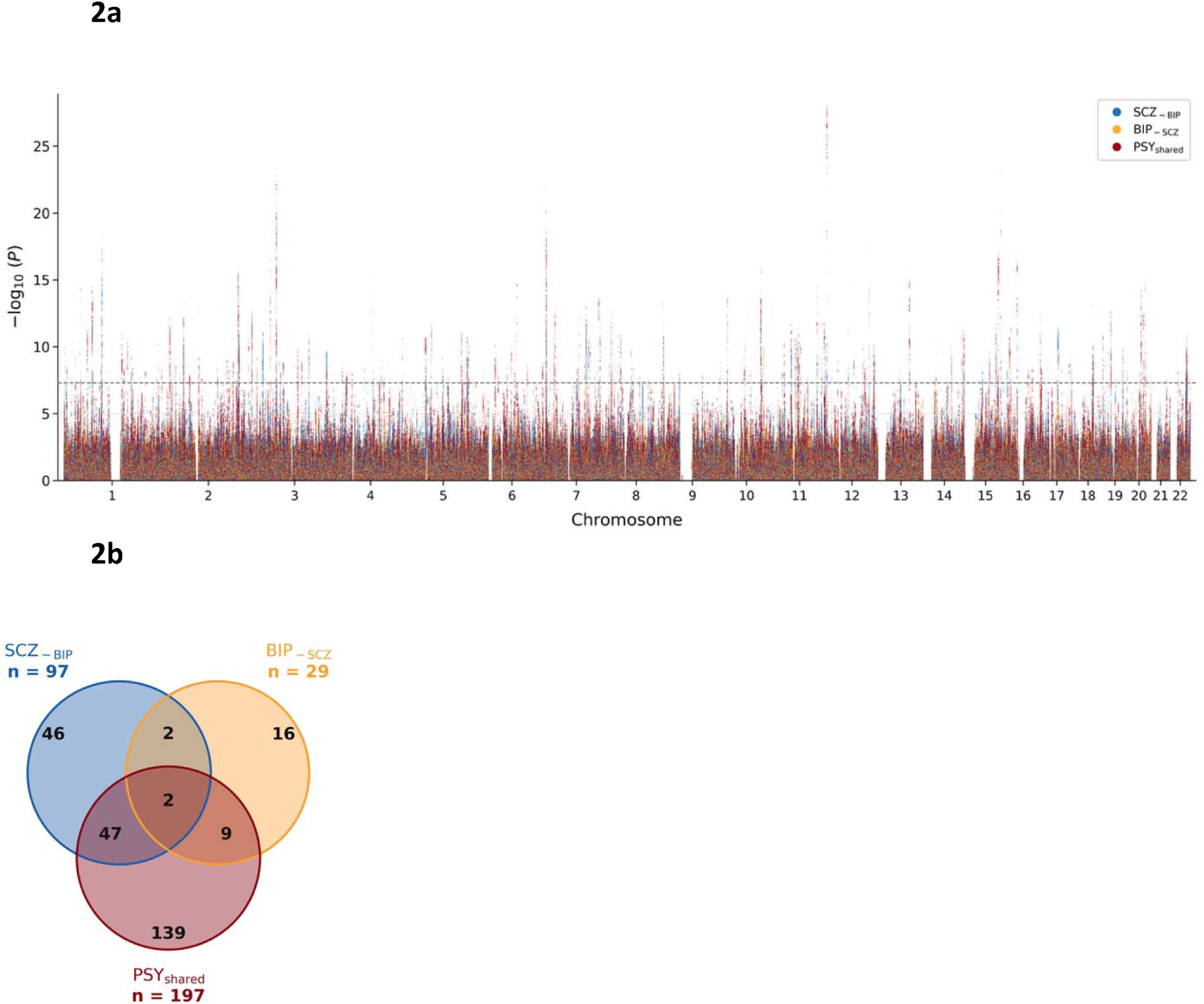
Genomic risk-locus landscape across the three components. (a) Manhattan plot of −log₁₀(P) for SCZ_∼BIP_, BIP_∼SCZ_, and Psy_shared_; the dashed line marks genome-wide significance (P = 5 × 10⁻⁸). (b) Venn diagram of 261 consensus loci obtained by merging component loci (SCZ_∼BIP_, n = 98; BIP_∼SCZ_, n = 29; Psy_shared_, n = 197) at ≥1-bp overlap on the same chromosome. Most loci were component-specific, and only four were shared between the orthogonal components.

### Novel component loci relative to the input-GWAS locus sets

A total of 80 loci identified in the derived components (64 PSY_shared_, 10 SCZ_∼BIP_, and 6 BIP_∼SCZ_) were not genome-wide significant in either input GWAS (Supplementary Table 2). We evaluated these loci against published multi-ancestry schizophrenia and bipolar-disorder GWASs^14–16,45–49^ and Genomic SEM psychosis-factor loci^8,9^, which showed that 64 (80%) had been reported previously. Sixteen loci remained novel relative to every comparison set: 14 PSY_shared_, one SCZ_∼BIP_, and one BIP_∼SCZ_. Of the 197 PSY_shared_ loci, 115 (58%) overlapped loci previously identified for the Genomic SEM schizophrenia-bipolar factor. A similar proportion of SCZ_∼BIP_ loci (56 of 98; 57%) also overlapped with loci previously identified for the Genomic SEM schizophrenia-bipolar factor, whereas the corresponding overlap was lower for BIP_∼SCZ_ loci (11 of 29; 38%).

### Gene-set enrichment (MAGMA and SynGO)

Using MAGMA, a total of 88 gene sets were FDR-significant for PSY_shared_, 58 for SCZ_∼BIP_, and two for BIP_∼SCZ_, compared with 78 for SCZ and 28 for BIP in the original input GWASs (Supplementary Table 3). Relative to the original SCZ GWAS, SCZ_∼BIP_ retained neuronal and synaptic enrichment, including gene sets such as dendritic tree (P_FDR_ = 1.27 × 10⁻⁵). Gene sets newly significant for SCZ_∼BIP_ included a range of biological pathways, such as cerebral-cortex GABAergic interneuron differentiation, beta-catenin-TCF complex, positive regulation of RNA metabolic process, and membrane depolarization. Of 34 SCZ-significant sets not retained by SCZ_∼BIP_, 24 were significant for PSY_shared_, including GABAergic and glutamatergic synapse, calcium-channel complex, and synapse organization. Notably, 26 gene sets were specific to SCZ_∼BIP_ that were not significant in PSY_shared_ (P_FDR_>0.05) and BIP_∼SCZ_ (P_FDR_>0.05 and nominal P > 0.001). These sets implicated both calcium and potassium ion transport across the membrane, as well as GABAergic interneuron differentiation, with additional annotations related to stress response, immune signaling, and muscle/cardiac function (Supplementary Table 3).

For BIP_∼SCZ_, only synapse (P_FDR_ = 4.31 × 10⁻²) and synaptic membrane (P_FDR_ = 4.69 × 10⁻²) survived correction. To characterize the weaker BIP_∼SCZ_ pathway signal, we additionally used nominal P < 0.001 as an exploratory criterion. Leading nominal associations involved postsynapse, neuronal-projection development, trans-synaptic signalling, axon extension, synaptic-vesicle priming and exocytosis, neurogenesis, and synaptic plasticity, but these did not reach corrected pathway-level significance. Of nominally significant gene sets for BIP_∼SCZ_, 19 gene sets were found to be specific to BIP_∼SCZ_ that were not nominally significant (p > 0.001) in SCZ_∼BIP_ and PSY_shared_. These sets included neuronal gene sets such as axon extension and synaptic-vesicle exocytosis, as well as more basic processes such as mitochondrial tRNA aminoacylation, histone ubiquitin-ligase activity, protein SUMOylation, and regulation of transcription-factor activity (Supplementary Table 3).

PSY_shared_ showed the broadest gene set enrichment profile, including synapse (P_FDR_ = 1.81 × 10⁻⁸), postsynaptic specialization (P_FDR_ = 4.38 × 10⁻⁸), high-voltage-gated calcium-channel activity (P_FDR_ = 4.38 × 10⁻⁸), and subpallium development (P_FDR_ = 7.32 × 10⁻⁶). It shared 55 significant sets with the input SCZ GWAS and 24 with input BIP, while 31 were significant newly in PSY_shared_. For example, gene set annotations for GABAergic synapse and glutamatergic synapse were significant in one or both input GWAS, and both were FDR-significant for PSY_shared_, but neither term reached FDR significance in either orthogonalized component. Similar results were also obtained for multiple ion channel annotations. Fifty-two gene sets were found to be specific to PSY_shared_ that were not significant for SCZ_∼BIP_ (P_FDR_ > 0.05) and BIP_∼SCZ_ (P > 0.001).

Several neurodevelopmental gene sets were specific to PSY_shared_ relative to the orthogonalized components, including striatum development, neuron development, and dendrite development. Surprisingly, a gene set related to neurodegeneration (tau protein kinase activity), was also significant for PSY_shared_ despite not previously being associated with SCZ or BIP (Figure 3; Supplementary Table 3). At the same time, 22 sets were significant in both PSY_shared_ and SCZ_∼BIP_, but not BIP_∼SCZ_ (even at the exploratory threshold; these largely reflected synaptic structure, neuronal development, and electrical signaling (Figure 3; Supplementary Table 3).

**Figure 3.**
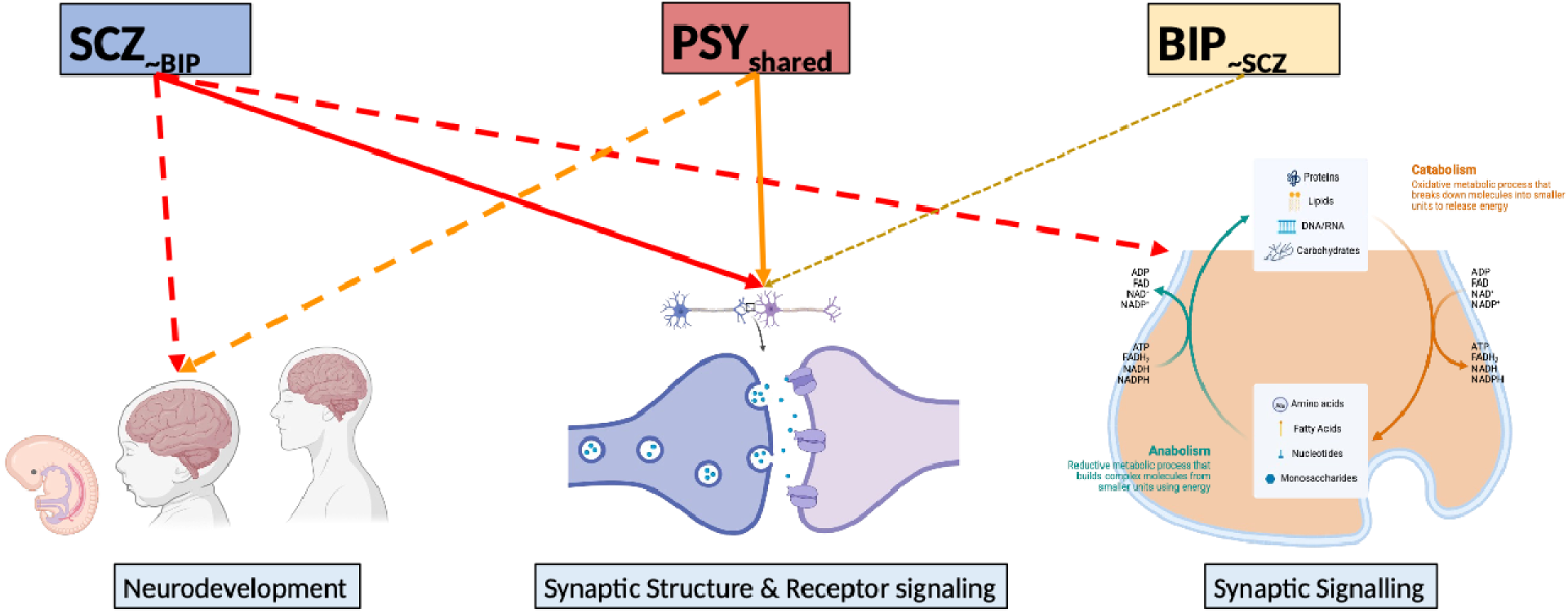
Biological pathway enrichment across SCZ_∼BIP_, BIP_∼SCZ_, and Psy_shared_. MAGMA competitive gene-set enrichment across the three components. Solid lines denote major enriched pathway groups, dashed lines additional groups with fewer enrichments, and dotted lines groups that did not reach FDR significance.

SynGO^44^ analysis refined the synaptic signal identified by MAGMA, identifying 32 significant annotations for SCZ_∼BIP_, 23 for BIP_∼SCZ_, and 29 for PSYshared (Supplementary Table 4). Although the profiles overlapped substantially, the additional SCZ_∼BIP_ annotations converged on postsynaptic organization and structural remodeling, whereas those for BIP_∼SCZ_ primarily involved synaptic membrane organization, vesicle release, and dendritic transport. PSY_shared_ largely captured features shared with both orthogonal components, showing greater overlap with SCZ_∼BIP_ in postsynaptic organization and with BIP_∼SCZ_ in synaptic membrane structure, but little enrichment unique to the shared component (Supplementary Table 4).

### Developmental timing of gene expression (BrainSpan)

BrainSpan gene-property analysis identified postnatal enrichment for SCZ_∼BIP_ in late infancy and young adulthood (both P_FDR_ = 3.35 × 10⁻³), while BIP_∼SCZ_ was significantly enriched only in late infancy (P_FDR_ = 3.30 × 10⁻²). By contrast, PSY_shared_ showed a strong prenatal enrichment (early-mid prenatal, P_FDR_ = 1.35 × 10⁻³; late-mid prenatal, P_FDR_ = 1.42 × 10⁻³) in addition to postnatal enrichments in late infancy (P_FDR_ = 1.80 × 10⁻⁵), and young adulthood (P_FDR_ = 1.35 × 10⁻³). This distinction is noteworthy, given that the input GWAS for both SCZ and BIP showed enrichment in both prenatal and postnatal gene expression in the brain (Supplementary Table 5).

### Genetic correlations among derived components

As intended, SCZ_∼BIP_ and BIP_∼SCZ_ were empirically uncorrelated (r_g_ = −0.025, SE = 0.029, P = 0.39), and both correlated positively with PSY_shared_ (SCZ_∼BIP_, r_g_ = 0.77; BIP_∼SCZ_, r_g_ = 0.62). SCZ_∼BIP_ correlated strongly with SCZ (r_g_ = 0.95) and moderately with BIP (r_g_ = 0.41); BIP_∼SCZ_ correlated strongly with BIP (r_g_ = 0.89) and weakly with SCZ (r_g_ = 0.26). PSY_shared_ correlated strongly with both SCZ (rg = 0.91) and BIP (rg = 0.89) (Figure 4a; Supplementary Table 6). ZCA therefore made the derived profiles orthogonal to each other, but not fully independent of the alternate input disorder; complete removal of the alternate-disorder correlation would make the two profiles strongly negatively correlated.

**Figure 4.**
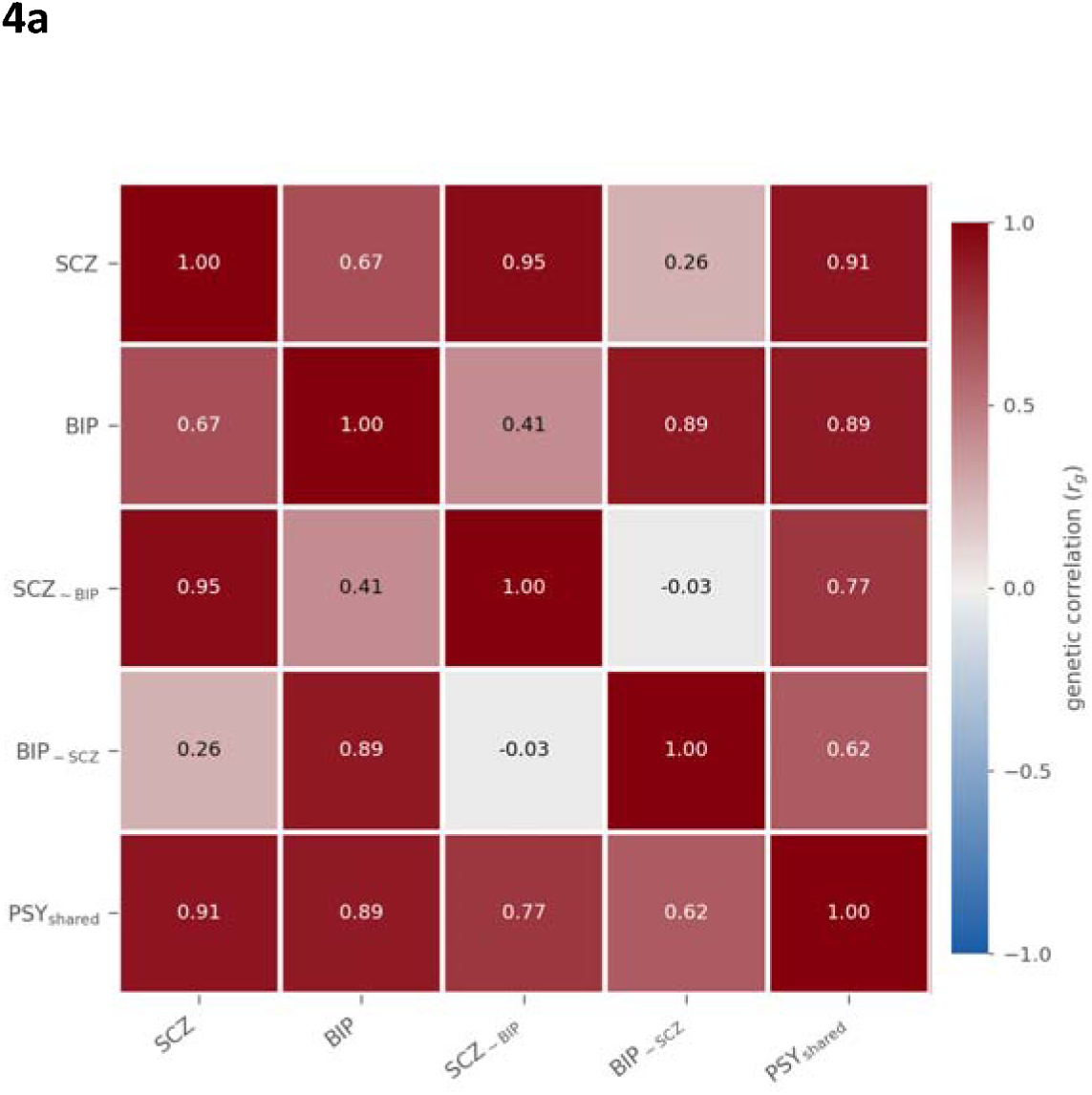
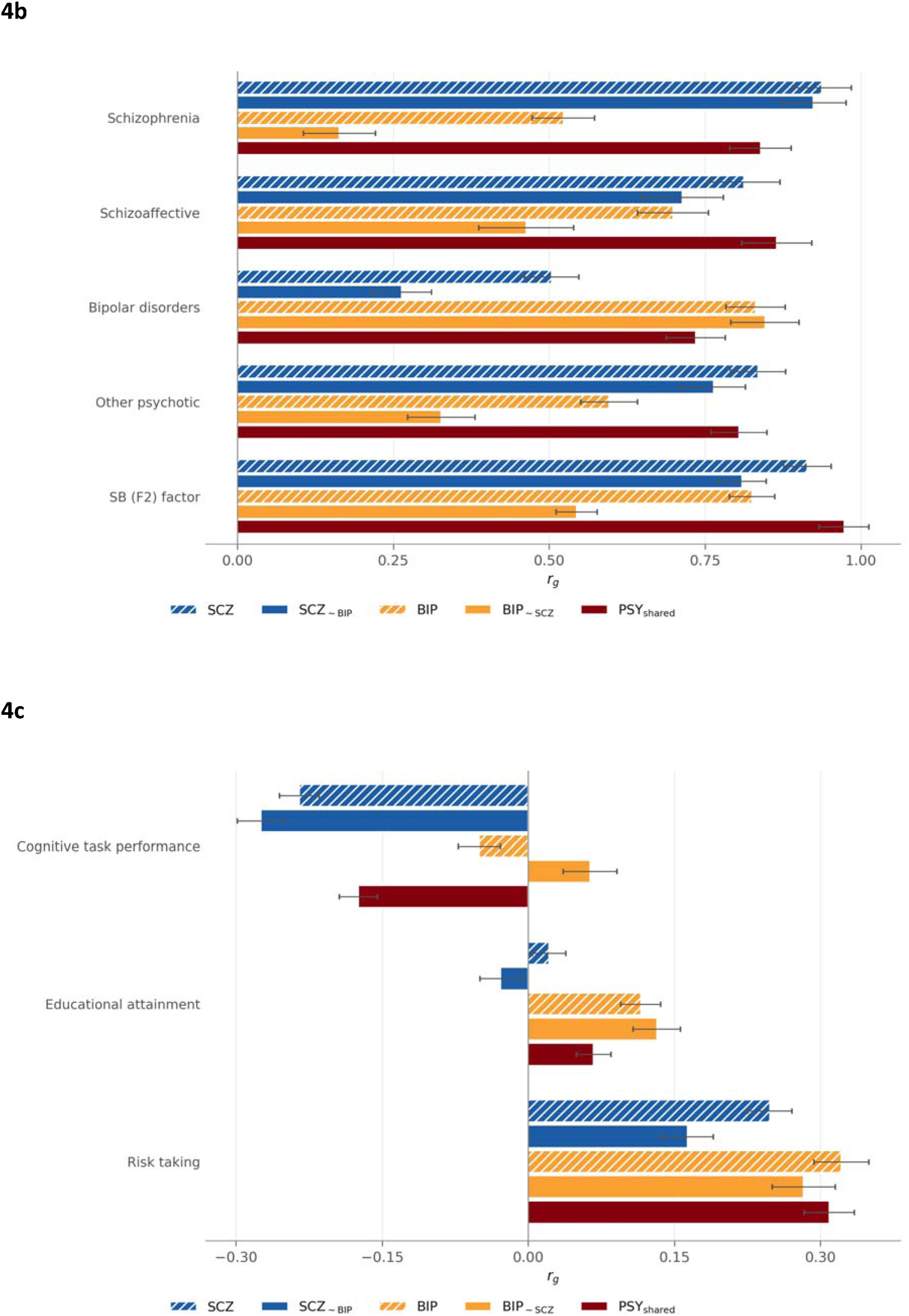
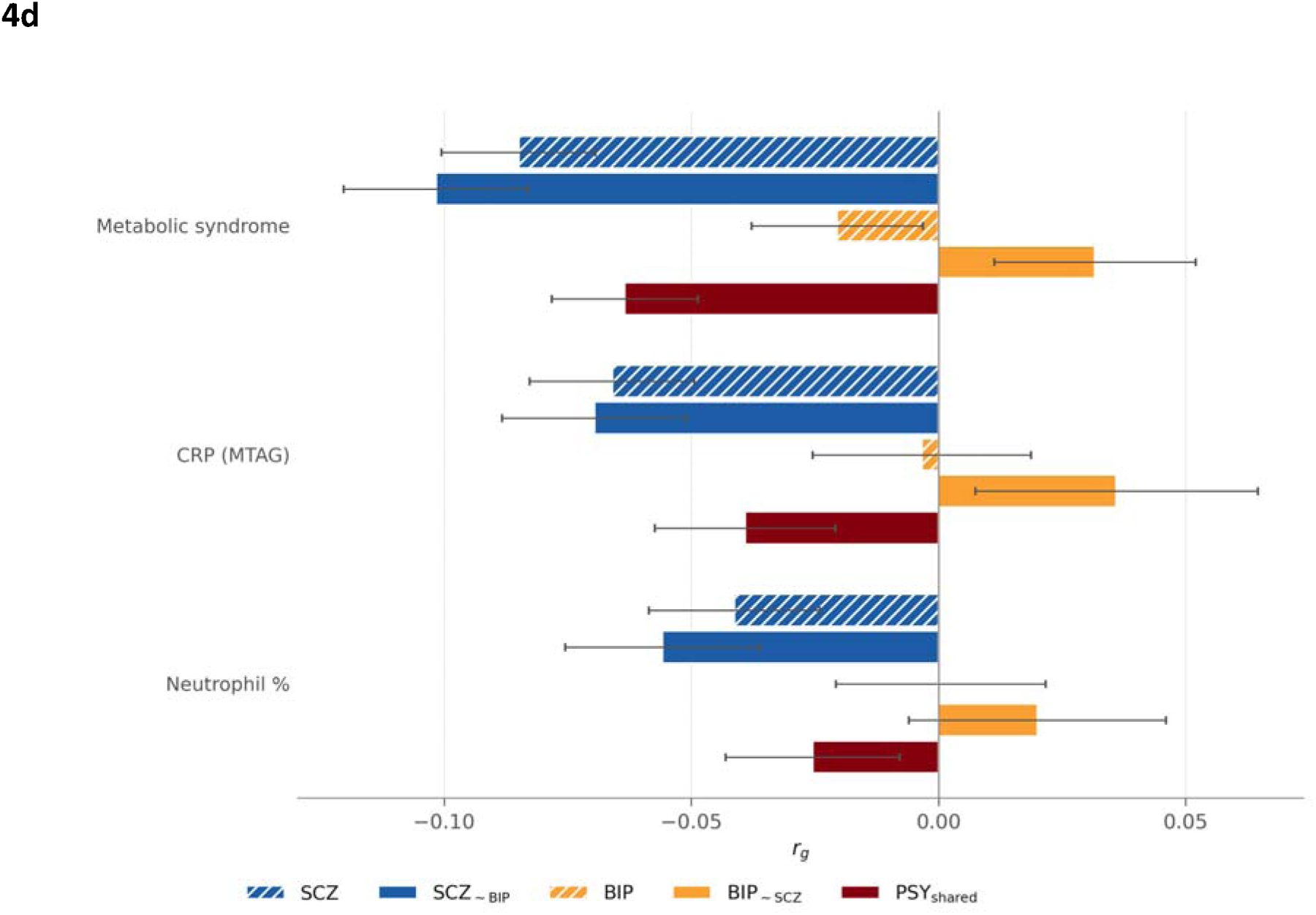
Genetic-correlation profiles of SCZ_∼BIP_, BIP_∼SCZ_, and Psy_shared_ relative to the original SCZ and BIP GWASs. LDSC genetic correlations (rg) are shown for the original disorders, the two ZCA components, Psy_shared_, and psychiatric, cognitive, cardiometabolic, and inflammatory reference phenotypes. Error bars in b-d denote ±1 standard error. a, Pairwise rg among SCZ, BIP, SCZ_∼BIP_, BIP_∼SCZ_, and Psy_shared_. The colour scale ranges from blue (rg = −1) to dark red (rg = +1); the diagonal is fixed at 1.0. The orthogonal components were empirically uncorrelated (rg = −0.03, P = 0.39), while both correlated positively with Psy_shared_ (SCZ_∼BIP_, rg = 0.77; BIP_∼SCZ_, rg = 0.62) and remained aligned with their source disorders (rg = 0.95 and 0.89). b, Genetic correlations with FinnGen psychosis-spectrum endpoints and the 14-disorder schizophrenia-bipolar factor. SCZ_∼BIP_ aligned most strongly with schizophrenia and other psychotic disorders, BIP_∼SCZ_ with bipolar disorder, and Psy_shared_ broadly across endpoints. Psy_shared_ showed the strongest correlation with the published schizophrenia-bipolar factor (rg = 0.97). c, Genetic correlations with cognitive-task performance, educational attainment, and risk taking. SCZ_∼BIP_ showed negative cognitive and educational estimates, BIP_∼SCZ_ positive estimates, and Psy_shared_ the mixed pattern seen in the input GWASs. All three components correlated positively with risk taking. d, Genetic correlations with metabolic syndrome, C-reactive protein, and neutrophil percentage. SCZ_∼BIP_ was significantly negatively correlated with all three phenotypes; BIP_∼SCZ_ estimates were positive but nonsignificant, and Psy_shared_ estimates were intermediate.

### Correlations with FinnGen psychiatric endpoints and the schizophrenia-bipolar factor

Independent FinnGen endpoints further distinguished the components. SCZ_∼BIP_ correlated most strongly with schizophrenia (r_g_ = 0.92), other and unspecified nonorganic psychosis (rg = 0.76), and schizoaffective disorder (r_g_ = 0.71), and much more weakly with bipolar disorder (r_g_ = 0.26) (Figure 4b; Supplementary Table 6). BIP_∼SCZ_ showed the reverse pattern, correlating most strongly with bipolar disorder (r_g_ = 0.85), followed by schizoaffective disorder (r_g_ = 0.46), other psychosis (r_g_ = 0.33), and schizophrenia (r_g_ = 0.16). As expected, PSY_shared_ correlated strongly across all FinnGen diagnoses: schizoaffective disorder (r_g_ = 0.86), schizophrenia (r_g_ = 0.84), other psychosis (r_g_ = 0.80), and bipolar disorder (r_g_ = 0.73). All three components correlated positively with the schizophrenia-bipolar factor from the 14-disorder Genomic SEM model^9^, most strongly for PSY_shared_ (r_g_ = 0.97), followed by SCZ_∼BIP_ (r_g_ = 0.81) and BIP_∼SCZ_ (r_g_ = 0.54) (Figure 4b; Supplementary Table 6).

### Genetic correlations with intermediate phenotypes

SCZ showed a significant negative correlation with cognitive-task performance^51^ (r_g_ = −0.27, SE = 0.024, P = 2.5 × 10⁻²⁹), and showed a negative (but non-significant) correlation with educational attainment^52^ (r_g_ = −0.028, P = 0.18). In contrast, BIP was positively correlated with cognitive performance (r_g_ = 0.063, P = 0.022) and educational attainment (rg = 0.13, P = 5.9 × 10⁻⁸). PSY_shared_ showed a mixed pattern: a significant negative correlation with cognitive performance (r_g_ = −0.17, P = 6.2 × 10^⁻19^) and a significant positive correlation with educational attainment (r_g_ = 0.067, P = 1.6 x 10^-4^) (Figure 4c; Supplementary Table 7). All three components showed significant positive correlations with risk taking: SCZ_∼BIP_ (r_g_ = 0.164, P = 6.61 × 10⁻¹⁰), BIP_∼SCZ_ (r_g_ = 0.283, P = 2.85 × 10⁻¹⁸) and PSY_shared_ (r_g_ = 0.309, P = 1.4 × 10⁻³³) (Figure 4c; Supplementary Table 7).

SCZ_∼BIP_ was also negatively correlated with metabolic syndrome (r_g_= −0.10, P = 5.20 × 10⁻⁸), C-reactive protein (r_g_ = −0.070, P = 2.01 × 10⁻⁴), and neutrophil percentage (r_g_ = −0.056, P = 4.58 × 10⁻³). By contrast, BIP_∼SCZ_ estimates were positive but nonsignificant for metabolic syndrome (r_g_ = 0.032, P = 0.12), C-reactive protein (r_g_ = 0.036, P = 0.21) and neutrophil percentage (r_g_ = 0.020, P = 0.44). PSY_shared_ estimates were intermediate, with significant negative correlations with metabolic syndrome (r_g_ = −0.063, P = 1.94 × 10⁻⁵), C-reactive protein (r_g_ = −0.039, P = 0.032), and a negative but non-significant correlation with neutrophil percentage (rg = −0.025, P = 0.15) (Figure 4d; Supplementary Table 7). As shown in Figure 4, the input SCZ correlations with all three traits were negative, whereas BIP estimates were near zero.

### Steiger tests for correlated correlations

Steiger tests^57^ showed that SCZ and BIP differed for all six intermediate phenotypes: cognitive-task performance (Δr_g_ = −0.338, P = 8.4 × 10⁻¹⁷), educational attainment (Δr_g_ = −0.160, P = 1.7 × 10⁻⁵), risk taking (Δr_g_ = −0.119, P = 9.2 × 10⁻³), metabolic syndrome (Δr_g_ = −0.133, P = 1.5 × 10⁻⁵), C-reactive protein (Δr_g_ = −0.106, P = 5.1 × 10⁻³), and neutrophil percentage (Δr_g_ = −0.076, P = 3.9 × 10⁻²). The corresponding differences for the input disorder-based GWAS were consistently smaller (Supplementary Table 7). Differences between the orthogonalized components exceeded the (input) disorder-level differences by 1.63-2.07-fold. A covariance-aware Wald test demonstrated that the correlated-correlation estimate difference between SCZ vs BIP and SCZ_∼BIP_ vs BIP_∼SCZ_ is significant for cognitive-task performance and metabolic syndrome (Supplementary Methods; Supplementary Table 8).

## Discussion

The present study is the first to apply ZCA (combined with Genomic SEM) to deconstruct GWAS data from schizophrenia and bipolar disorder. Our framework is inspired by the longstanding evidence of phenotypic^10^ and genetic^7,8,9^ overlap between schizophrenia and bipolar disorder, as well as the recently proposed stratagenic model^17^, which posits that genetic risk may be distributed across functionally distinguishable biological processes. This framework enabled us to evaluate whether decomposition revealed differentiated sets of pathways and associations that were less apparent in the aggregate SCZ and BIP GWASs.

Of 261 consensus loci identified in our study, 201 (77%) were associated with only one component. Moreover, we identified 80 loci that were not genome-wide significant in either input GWAS. Comparison against all published schizophrenia and bipolar-disorder GWASs^14–16,45–49^ and both Genomic SEM psychosis-factor locus sets^8,9^ showed that 64 of these 80 loci (80%) had been reported in at least one study, indicating that our decomposition retrieves genuine signal despite smaller sample size relative to the most recent transancestry GWAS. An additional 16 loci were entirely novel with respect to the larger SCZ and BIP studies available to date. Alignment with independent clinical endpoints from FinnGen supported the intended interpretation of the components. Because the FinnGen endpoints are registry-based and derived from an independent population, this alignment is not an artefact of the input summary statistics used for decomposition.

The gene set enrichment results extend the same observation to biology; while there was a degree of overlap, with all three components demonstrating neuronal/synaptic gene set enrichments, there were sufficient differences to yield distinguishable biological profiles. PSY_shared_ showed the broadest enrichment, spanning synaptic compartments, postsynaptic organisation, plasticity, glutamatergic and GABAergic synaptic architecture, vesicle trafficking and ion transport, consistent with a substantial neuronal substrate common to both diagnoses. Gene sets significant only in PSY_shared_ included nuclear body and nuclear speck, suggesting involvement of the spatial organization of transcription and RNA processing. Nuclear speckles concentrate RNA-processing machinery, and proximity to these compartments increases the efficiency of pre-mRNA splicing^58^. In developing human cortex, repositioning neuronal genes towards nuclear speckles facilitates their transcriptional activation, connecting nuclear organization to coordinated neuronal gene-expression programmes^59^. Alterations in cortical RNA splicing have been reported in schizophrenia and bipolar disorder, with differentially spliced genes across the psychiatric disorders studied involving synaptic, signaling and neuronal-development functions^60^.

Several neurodevelopmental gene sets were also specific to the PSY_shared_ component, consistent with evidence that the two disorders share a neurodevelopmental substrate^61^, and contrary to the hypothesis that neurodevelopmental anomalies are specific to schizophrenia^62^. Evidence for an early developmental contribution has historically been stronger for SCZ than for BIP^61,63^, although epidemiological, neuroimaging, and postmortem data also support neurodevelopmental contributions to BIP^63,64^. Temporal expression analysis using BrainSpan data yielded comparable results. Before decomposition, both input GWASs showed enrichment across prenatal and postnatal windows; after decomposition, prenatal enrichment reached significance only in PSY_shared._ SCZ_∼BIP_ and BIP_∼SCZ_ were both significantly enriched in genes expressed in late infancy, while SCZ_∼BIP_ was also enriched in young adulthood. These profiles are compatible with their respective gene set enrichment results elaborated immediately below, in the context of the changing transcriptional programs of the developing human brain^65,66^.

SCZ_∼BIP_ showed enrichment for cortical differentiation annotations, including cerebral-cortex GABAergic-interneuron differentiation and cortical-neuron differentiation, that met significance only in that component. These findings are compatible with evidence of altered GAD67 and parvalbumin expression in cortical interneurons in schizophrenia and models connecting inhibitory-circuit dysfunction to working memory and gamma oscillations^67,68^. The DNA-and RNA-related enrichments in SCZ_∼BIP_ extend its biological profile beyond neuronal development. SCZ_∼BIP_-specific pathways involving protein-DNA complex, AT-rich DNA binding, transcription-initiation-factor binding and beta-catenin-TCF complex likewise point to multiple aspects of transcriptional control. Chromatin organization and transcriptional regulation coordinate neuronal gene programs by controlling access to regulatory DNA and interactions between enhancers and promoters. In developing human cortex, chromatin-contact maps linked schizophrenia-associated variants to candidate pathways involving neurogenesis and cholinergic signalling^69^. Postmortem midbrain analyses identified schizophrenia-risk-enriched downregulated genes within dopamine-neuron-specific chromosomal domains^70^. Our earlier investigation of the genetic relationship between schizophrenia and cognition^25^ similarly implicated various DNA/RNA and chromosomal organization pathways as observed in the current study as well. These findings suggest that DNA-binding and transcriptional alterations may reflect coordinated regulation of neuronal functions specific to schizophrenia and directly related to the underlying cognitive deficits in that disorder.

By contrast, BIP_∼SCZ_ only retained FDR-corrected synapse and synaptic-membrane enrichment, with additional projection-development, plasticity and vesicle-release associations apparent at the nominal (P < 0.001) threshold. At this exploratory threshold, gene sets specific to BIP_∼SCZ_ included mitochondrial tRNA aminoacylation, consistent with recent evidence suggesting a role for mitochondrial dysfunction in the etiology of bipolar disorder^71^. Aminoacylation supplies charged tRNAs for mitochondrial translation^72^, linking this annotation to maintenance of the machinery that supports oxidative energy production. Deep-exome sequencing of peripheral tissues identified enrichment of heteroplasmic mitochondrial tRNA variants in bipolar disorder, providing indirect genetic support for mitochondrial RNA biology^73^. Other recent studies provide evidence that lithium may act to restore mitochondrial function^74^, suggesting that alternative strategies such as ketogenic diet may have therapeutic effect^75^.

The intermediate-phenotype analyses bear most directly on whether decomposition informs clinical heterogeneity. Diagnostic categories alone do not account for the substantial variation in cognition^12,76^, cardiometabolic burden^28,30,77^, and inflammatory status^35,78^ observed within SCZ and BIP, and cognitive clustering studies have repeatedly identified subgroups that cross diagnostic boundaries, including globally impaired BIP groups^79^ whose profiles resemble those more typically observed in SCZ^80^. For the original, diagnosis-based GWASs, genetic relationships with these phenotypes were asymmetric: SCZ was negatively correlated with cognitive performance, metabolic syndrome, CRP, and neutrophil percentage, whereas the corresponding BIP estimates were near zero. After decomposition, SCZ_∼BIP_ and BIP_∼SCZ_ diverged in opposite directions across cognitive-task performance, educational attainment, metabolic syndrome, CRP and neutrophil percentage. The component contrast exceeded the disorder-level contrast by a factor of 1.63-2.07, with the increase remaining significant after correction for cognitive task performance and metabolic syndrome. At the same time, the PSY_shared_ component correlated with these most of these phenotypes in a pattern similar to, though to a lesser degree, SCZ_∼BIP_, with the notable exception of educational attainment. Thus, individuals with the same DSM-5 diagnosis may differ in their relative burden of the three liability components, offering a genetic account of variation along the cognitive, metabolic, and inflammatory spectra observed clinically. The pattern of associations with risk taking were somewhat surprising: rather than a specific association with BIP_∼SCZ_, all three components were positively correlated with risk taking, with PSY_shared_ showing the strongest correlation.

Prior multivariate analyses of correlated psychiatric GWASs, though asking different questions from the one posed here, established the methodological foundation for the present work. Genomic SEM was developed as a general framework for modelling the genetic covariance structure among GWAS summary statistics^20^, where SCZ and BIP were consistently recovered as indicators of a single factor^8,9^, without further resolving structure within this factor. Recently, Watson et al. applied GWAS-by-subtraction^81^ to schizophrenia and bipolar disorder GWAS summary statistics, specifying a SZ-specific factor that captured the genetic variance in schizophrenia not accounted for by the liability shared with bipolar disorder. Because the model was designed to partition schizophrenia liability, rather than to derive parallel residual components for both disorders, it did not estimate a BIP-specific or BIP-predominant dimension^82^. Furthermore, mtCOJO^83^ can provide a complementary conditional perspective, adjusting each psychiatric-disorder GWAS for genetically correlated disorders to identify putatively disorder-specific SNP associations. Although this method has been used to generate conditional SCZ and BIP GWAS results, its focus was locus-level specificity across disorders; the smaller BIP GWAS also constrained power for BIP-specific discovery^21^. In the present study, mtCOJO results were comparable, though statistically weaker, than results provided by ZCA (see Supplement). CC-GWAS represents an important advance for identifying genetic differences between psychiatric diagnoses. However, because it is a pairwise case-case comparison, it estimates loci that differentiate diagnostic groups^22^ rather than generating separable genome-wide dimensions of SCZ-predominant and BIP-predominant liability, or assessing their relative contributions to heterogeneity within either diagnosis. Taken together, these studies show that psychiatric risk can be characterized in terms of shared liability, disorder-conditioned associations, and genetic differences between diagnostic groups. However, no prior study was designed to test how disease heterogeneity might arise from the relative contributions of shared, SCZ-predominant, and BIP-predominant genetic liability.

Across levels of analysis, the three components showed patterns that were not apparent in the aggregate disorder GWASs. Most consensus loci were associated with only one component, and functional analyses identified a common synaptic substrate alongside differently weighted cortical differentiation, gene-regulatory, protein-turnover, mitochondrial and secretory pathways. Developmental expression enrichment further distinguished the shared and disorder-predominant profiles. Most importantly, SCZ_∼BIP_ and BIP_∼SCZ_ showed opposing associations with cognitive, cardiometabolic, and inflammatory phenotypes, whereas the corresponding disorder-level associations were predominantly asymmetric or attenuated. Thus, the decomposition did more than redistribute genetic signal across derived GWASs: it exposed differentiated patterns of association that are obscured when shared and disorder-predominant influences are combined within diagnostic GWAS statistics. These results are consistent with the possibility that individuals assigned the same diagnosis differ in the relative weighting of shared, SCZ-predominant, and BIP-predominant liability. The present analyses cannot establish clinical subtypes or individual-level prediction, but they provide a genetically informed framework for testing whether this multidimensional liability profile accounts for variation in cognition, cardiometabolic and inflammatory burden, illness course, or treatment response in deeply phenotyped cohorts.

## Limitations

The analyses used European-ancestry GWASs, so generalizability to other ancestry groups remains to be evaluated. The parent BIP GWAS and the derived BIP_∼SCZ_ component had lower liability-scale SNP heritability than their SCZ counterparts, and the published European-ancestry BIP analysis contained substantially fewer genome-wide significant loci. These differences reduce the breadth of locus-and pathway-level findings available for BIP_∼SCZ_ and make the absence of additional corrected enrichments less informative. This asymmetry is not attributable to a smaller effective sample size and should not be interpreted as failure of the decomposition. ZCA makes the two derived components mutually orthogonal but does not make either independent of the alternate original disorder; they should therefore be interpreted as disorder-predominant rather than disorder-specific. Because the components are algebraic functions of the input GWASs, their exact composition is input-dependent. The six external phenotypes were hypothesis-guided anchors rather than an exhaustive or independent phenome-wide validation set.

## Supporting information

Supplementary Methods and Result

## Data Availability

All data produced in the present study are available upon reasonable request to the authors

