## Supplementary Methods and Result for "Disorder-specific and shared genetic architecture underlying schizophrenia and bipolar disorder"

### ^1^Northwell, New Hyde Park, NY, USA.

### ^2^Institute of Behavioral Science, Feinstein Institutes for Medical Research, Manhasset, NY, USA.

### ^3^Institute of Mental Health, Singapore

### ^4^Lee Kong Chian School of Medicine, Population and Global Health, Nanyang Technological University

### ^5^Departments of Psychiatry and Molecular Medicine, Zucker School of Medicine at Hofstra/Northwell, Hempstead, NY

### *These authors contributed equally

### §Corresponding Author:

### Todd Lencz; phone: +1 917-453-4647;; Office: Division of Psychiatry Research, Zucker Hillside Hospital, Glen Oaks, NY, USA 11004

#### Supplementary Methods and Results

##### GWAS summary statistics

GWAS summary statistics for schizophrenia were obtained from the Psychiatric Genomics Consortium third-wave meta-analysis (PGC3; 53,386 cases and 77,258 controls)^1^, and bipolar-disorder statistics from the PGC bipolar disorder working group (59,287 cases and 781,022 controls)^2^. FinnGen release R13 endpoints spanning schizophrenia, bipolar disorder, schizoaffective disorder and other psychotic disorders^3^, and the schizophrenia–bipolar latent factor from the 14-disorder Genomic SEM analysis^4^, were used for external characterisation.

To characterize each derived component against clinically relevant intermediate phenotypes and comorbidities, we obtained European-ancestry GWAS summary statistics for cognitive task performance (CTP; N = 373,617)^5^, educational attainment (EDU; N = 766,345)^6^, risk taking (UK Biobank field 2040; 104,809 cases and 301,022 controls)^7^, metabolic syndrome (MetS; N = 1,384,348)^8^, C-reactive protein (CRP; N = 575,531)^9^ and neutrophil percentage of leukocytes (N = 394,642)^10^. Risk-taking statistics were taken from the Pan-UK Biobank European-ancestry analysis and harmonised through the pipeline described below.

##### Quality control and harmonisation

Each GWAS summary-statistic dataset was subjected to a standardised quality-control pipeline as described previously^11^. Summary statistics were harmonised using gwas2vcf^12,13^, which converts GWAS results to Variant Call Format, standardises allele representation to GRCh37 and aligns reported effect directions. Variants with zero effect-size estimates or standard errors were removed.

We also removed indels, strand-ambiguous A/T and C/G SNPs, variants with INFO ≤ 0.3, variants with MAF ≤ 0.01, and variants in the extended MHC region (chromosome 6: 25,000,000–35,000,000 bp, GRCh37). The 1000 Genomes Project Phase 3 European panel (GRCh37) was used for LD estimation in LDSC, FUMA and MAGMA.

##### Derivation of the disorder-predominant components by ZCA whitening

###### Zero Component Whitening transformation and rationale

Zero-phase component (ZCA) whitening was applied to the bivariate SNP-based genetic covariance matrix for SCZ and BIP to derive two mutually orthogonal, disorder-predominant components. ZCA was selected because it is symmetric, order-invariant and minimises the expected squared displacement of the transformed variables from the inputs among whitening transformations^14^. ZCA whitening has previously been used to decorrelate multi-trait GWAS z-score vectors^15,16^. The components were interpreted as orthogonal profiles that remain closely aligned with their respective parent GWASs, rather than as complete residuals from the alternate disorder.

The liability-scale covariance matrix Σ was estimated by bivariate LDSC as implemented in Genomic SEM^17^, using HapMap3 variants and 1000 Genomes Project Phase 3 European LD scores. Sample prevalence was set to 0.5 with effective sample size, and population prevalence to 0.01 for SCZ and 0.02 for BIP. The matrix was Σ = [[0.212314, 0.092447], [0.092447, 0.090159]], and the cross-trait LDSC intercept used for sampling-covariance propagation was ρ = 0.1636.

ZCA whitening was defined by WΣWᵀ = I, with W = Σ⁻¹ᐟ². Eigendecomposition of Σ gave W = [[2.631053, −1.259673], [−1.259673, 4.295515]].

###### SNP-level construction

For variant j, aligned parent-GWAS effects were transformed as β*j = Wβj, where βj = (βSCZ,j, βBIP,j)ᵀ. Up to component-specific scaling, this is equivalent to SCZ_~BIP_ ∝ βSCZ − 0.478771βBIP and BIP_~SCZ_ ∝ βBIP − 0.293253βSCZ.

Because the input GWASs included overlapping samples, per-variant sampling variance was propagated using
ρ = 0.1636: Var(β*i,j) = wi1²SESCZ,j² + wi2²SEBIP,j² + 2wi1wi2ρSESCZ,jSEBIP,j.
Composite standard errors, Z statistics and P values were then derived from the transformed effects and variances.

###### Scale and orthogonality checks

The transformation gives each component unit genetic variance and zero genetic covariance with the other component by construction; it does not require either component to be uncorrelated with the alternate parent GWAS. Empirically, the two derived components were nearly uncorrelated by LDSC (rg = −0.025, SE = 0.029), consistent with the intended target.

As a focused relative-scale check, the composites were reconstructed in 50,000 deterministic matched SNPs using both the production beta/SE inputs and the corresponding Genomic SEM-standardised inputs. The resulting Z statistics were essentially identical (Pearson r > 0.999999999; maximum |ΔZ| ≤ 0.00142), supporting use of the reported whitening coefficients without additional rescaling.

##### Derivation of disorder-predominant components using mtCOJO

We recognize that ZCA whitening is a relatively rare method to be used in study design like ours, thus we also derived disorder-predominant profiles using multi-trait conditional and joint analysis (mtCOJO)^18^ implemented in GCTA v1.94.1. mtCOJO conditions the effect-size estimates of one trait on the genetically predicted contribution of another using GWAS summary statistics and an LD reference panel. The resulting profiles are conditional estimates; mtCOJO does not impose orthogonality.

Two conditional analyses were performed:

1. SCZ|BIP: schizophrenia GWAS summary statistics conditioned on the genetically predicted contribution of BIP, yielding a schizophrenia-predominant conditional profile.

2. BIP|SCZ: bipolar-disorder GWAS summary statistics conditioned on the genetically predicted contribution of SCZ, yielding a bipolar-predominant conditional profile.

Alleles were harmonised to GRCh37 before analysis. For both directions, LD was estimated using the 1000 Genomes Project Phase 3 European-ancestry GRCh37 panel (503 individuals; 22,238,705 variants). Default mtCOJO/GSMR settings were retained for instrument selection (P < 5 × 10⁻⁸), LD clumping (r² < 0.05), HEIDI-outlier filtering (P < 0.01), minimum instruments (10) and LD FDR (0.05); the analysis additionally specified an allele-frequency-difference threshold of 0.20 (--diff-freq 0.2) and a 5,000-kb conditional-analysis window (--cojo-wind 5000). Sample and population prevalences were 0.4086 and 0.01 for SCZ and 0.2114 and 0.02 for BIP. For SCZ|BIP, 74 BIP instruments were identified, six were removed by HEIDI and 68 were retained; the conditioning coefficient was 0.637683 (SE = 0.025531). For BIP|SCZ, 195 SCZ instruments were identified, five were removed and 190 were retained; the coefficient was 0.322468 (SE = 0.010277). Other settings remained at GCTA defaults.

##### Derivation of the SCZ–BIP shared component using Genomic SEM

The genetic component shared by SCZ and BIP (Psy_shared_) was estimated from the bivariate LDSC genetic and sampling covariance matrices using Genomic SEM^17^ v0.0.5 (CommonFactor GWAS), lavaan v0.6-21 and R v4.5.3. Harmonised inputs included SNP identifiers, alleles, Z statistics, P values and per-variant effective sample sizes and were processed using GenomicSEM::sumstats(linprob = TRUE, se.logit = FALSE).

A preliminary SNP-free calibration model fixed factor variance to one and estimated the indicator loadings and residual variances, returning unstandardised loadings of 0.338258 for SCZ and 0.272208 for BIP. These values were rounded and fixed at 0.34 and 0.27, respectively, in the final measurement model.

SNP effects on Psy_shared_ were estimated using the default diagonally weighted least-squares estimator in GenomicSEM::userGWAS(fix_measurement = TRUE), in 5,000-SNP chunks, while accounting for the genetic and sampling covariance of the parent GWASs.

##### Genomic risk-locus definition and candidate-gene mapping

Genome-wide significant variants (P < 5 × 10⁻⁸) from each component GWAS were processed using Functional Mapping and Annotation of Genetic Associations (FUMA) v2.1.5^19^ to define genomic risk loci and identify candidate genes.

**Risk-locus definition.** FUMA defines risk loci through a two-stage clumping procedure. First, independent significant SNPs were identified as variants reaching P < 5 × 10⁻⁸ that are independent of each other at a linkage-disequilibrium threshold of r² < 0.6. Second, among independent significant SNPs, lead SNPs were identified by further clumping at r² < 0.1. Genomic risk loci were then defined by merging lead SNPs (and their associated LD-defined regions) located within 250 kb of each other into single loci. The LD reference panel for clumping was the 1000 Genomes Project Phase 3 European-ancestry panel. The boundaries of each risk locus were defined by the most extreme positions of all candidate SNPs (those in LD with independent significant SNPs at r² ≥ 0.6 and with P < 0.05) within each locus. All locus counts reported in this study are LD-based FUMA counts defined in this way, not distance-based clumping counts.

**Candidate-gene mapping.** Within each defined risk locus, candidate genes were prioritised using three complementary mapping strategies implemented in FUMA: (i) *positional mapping*, in which genes were assigned to a risk locus if any part of the gene body fell within a 10-kb window of the locus boundaries; (ii) *expression quantitative trait locus (eQTL) mapping*, in which SNPs within risk loci were linked to genes whose expression they significantly regulate (at FDR < 0.05) across all tissue panels available in FUMA version 2.1.5, a SNP–gene pair being included if the SNP was a significant eQTL for that gene in at least one tissue; and (iii) *three-dimensional chromatin-interaction mapping*, in which SNPs were linked to genes through significant chromatin contacts identified in all Hi-C datasets available in FUMA version 2.1.5, an interaction being considered significant if it connected a genomic region containing a candidate SNP to a region overlapping a gene promoter (defined as 250 bp upstream and 500 bp downstream of the transcription start site) with FDR < 1 × 10⁻⁶. The union of genes identified across the three mapping strategies constituted the final candidate gene set for each component and was carried forward to all downstream functional analyses.

##### Locus-overlap and novelty analyses

We quantified locus sharing and distinctness across the three liability components using BEDTools v2.30.0^20^.

**Cross-component overlap.** The genome-wide significant risk loci defined by FUMA for each of the three components were compared pairwise and jointly. Overlap between two loci was defined as a minimum of one shared base position (1-bp overlap using BEDTools intersect). Loci were classified into: (i) consensus loci, the non-redundant union of all risk loci across the three components (obtained using BEDTools merge); (ii) loci shared across all three components; (iii) loci shared between exactly two components; and (iv) loci unique to a single component.

Novelty against input GWASs, published PGC studies and Genomic SEM SCZ–BIP factors. Component loci were first classified as absent in the input if they did not overlap a genome-wide significant locus from either input GWAS^1,2^. For a stricter novelty assessment, we also compared the loci identified with five published schizophrenia GWAS locus sets^1,21–24^, three published bipolar-disorder GWAS locus sets^2,25,26^, and SCZ–BIP factor loci from the 11- and 14-disorder Genomic SEM studies^4,27^. Overlap required at least one shared base on the same chromosome in GRCh37. Bigdeli et al. loci were lifted from GRCh38 to GRCh37 using liftOver; three intervals that failed liftOver were excluded. Publications where loci were not reported, we used a ±20-kb interval from the reported lead SNP to find overlap with loci identified in this study. Loci absent from every comparison set were classified as novel with respect any large scale PGC SCZ and BIP available till date; however, we understand that these loci could have been present in other meta-analysis or multivariate studies including SCZ and BIP (Supplementary Tables 1 and 2).

##### Competitive gene-set enrichment analysis (MAGMA)

Gene-set enrichment analysis was performed using Multi-marker Analysis of Genomic Annotation (MAGMA) v1.10^28^ to identify biological pathways and functional categories over-represented among genes implicated by each liability component.

**Gene annotation.** SNPs were annotated to 18,877 protein-coding genes (NCBI build 37, gene-boundary definitions from the NCBI gene database) using an asymmetric window extending 35 kb upstream and 10 kb downstream of each gene body. The upstream extension was chosen to capture promoter regions and proximal enhancers, while the downstream extension captures 3′ regulatory elements.

**Gene-based association testing.** For each of the three components, gene-level association statistics were computed using the MAGMA SNP-wise mean model, which combines the association P values of all SNPs annotated to a gene into a single gene-level test statistic. Inter-SNP linkage disequilibrium was modelled using the 1000 Genomes Project Phase 3 European panel to ensure that correlated SNPs within a gene do not inflate the gene-level statistic.

Competitive gene-set testing. Genes on sex chromosomes and within the extended MHC region (chromosome 6: 25–35 Mb) were removed from all gene sets, and gene sets containing fewer than five remaining genes were excluded. Gene-level statistics were tested against Gene Ontology Biological Process, Cellular Component and Molecular Function collections, Reactome pathways, and curated gene sets. Competitive models included gene size, gene density and inverse minor allele count as covariates. Benjamini–Hochberg FDR correction was applied separately within each component across all tested gene sets (Supplementary Table 3).

##### SynGO synaptic gene-set enrichment

Synapse-specific enrichment was evaluated using SynGO release 1.3^29^. Unique gene symbols with nominal MAGMA gene-based P < 0.05 were submitted for SCZ_~BIP_ (4,494 genes), BIP_~SCZ_ (3,409), Psy_shared_ (6,093), SCZ|BIP (4,046), BIP|SCZ (3,332), SCZ (6,051) and BIP (4,624). Over-representation analysis used the default built-in brain-expressed background, a minimum of three genes per term and one-sided Fisher exact tests (Supplementary Table 5).

##### BrainSpan developmental age

To determine whether genes implicated by each component were preferentially expressed during specific developmental periods, we performed MAGMA gene-property analysis using transcriptomic data from the BrainSpan Atlas of the Developing Human Brain^30^ implemented within FUMA. BrainSpan RNA-sequencing profiles span 31 developmental stages from early prenatal life to late adulthood, collapsed here into eleven developmental periods.

For each component, the MAGMA gene-property model related gene-level association statistics to average gene expression within each developmental period, conditional on average expression across all periods and the standard MAGMA gene-level covariates.

Multiple-testing correction was applied using the Benjamini–Hochberg procedure to control the FDR at 5% within each component, across that component’s eleven developmental periods.

##### Characterisation of the three-component decomposition

We assessed whether SCZ_~BIP_, BIP_~SCZ_ and Psy_shared_ showed the intended inter-component relationships and distinguishable associations with external psychiatric endpoints.

###### Inter-component genetic correlations

Pairwise genetic correlations among the three components were estimated using LDSC. The correlation between the ZCA disorder-predominant components fell from 0.67 in the parent GWASs to −0.025 (SE = 0.029), consistent with the orthogonality target, and both remained moderately correlated with Psy_shared_.

###### Genetic correlations with independent psychiatric endpoints

We estimated genetic correlations with four independent FinnGen release R13 psychiatric endpoints^3^ and the schizophrenia–bipolar latent factor from the 14-disorder Genomic SEM analysis^4^. These analyses assessed whether SCZ_~BIP_ aligned preferentially with schizophrenia-spectrum endpoints, BIP_~SCZ_ with bipolar disorder, and Psy_shared_ with endpoints representing liability common to SCZ and BIP.

All genetic correlations were estimated using LDSC^31^ with pre-computed 1000 Genomes Project Phase 3 European LD scores. Benjamini–Hochberg correction controlled the FDR at 5% across component–phenotype pairs (Supplementary Tables 6 and 7).

##### Characterization of genetic correlation profiles across endophenotypic domains

The ZCA components (SCZ_~BIP_, BIP_~SCZ_), mtCOJO components (SCZ|BIP, BIP|SCZ), input GWASs, Psy_shared_ and reference phenotypes were analysed jointly using GenomicSEM::ldsc() with a 200-block jackknife, yielding genetic covariance matrix S and sampling covariance matrix V. Joint analysis was required because the component and parent-GWAS estimates are dependent. Each LDSC estimate used the harmonised HapMap3 variants available for the relevant profiles.

Steiger tests for correlated genetic correlations^32^ compared rg(T,A) with rg(T,C) for each external phenotype T (CTP, EDU, risk taking, MetS, CRP and neutrophil percentage) within the ZCA, mtCOJO and parent-GWASs. Because each pair shared the same external phenotype, sampling covariance was obtained from the corresponding off-diagonal element of the 200-block LDSC jackknife covariance matrix V after delta-method rescaling. The statistic was Z = (rgA − rgC)/√(SEA² + SEC² − 2cov[rgA,rgC]), with two-sided P values. Jackknife estimates were primary; analytic Olkin/Steiger covariance^33^ was examined as a sensitivity analysis. Benjamini–Hochberg correction was applied across the six external traits.

###### Wald test for comparison of genetic-correlation contrasts after decomposition

To test whether the SCZ–BIP genetic-correlation contrast changed after decomposition, we compared Δcontrast = [rg(SCZ_~BIP_,X) − rg(BIP_~SCZ_,X)] − [rg(SCZ,X) − rg(BIP,X)] with zero for each external trait X; the corresponding analysis used SCZ|BIP and BIP|SCZ (mtCOJO derived component).

The four correlations and their full sampling covariance were obtained from S and V by the delta method, with the Jacobian evaluated using numDeriv v2016.8-1.1 (Richardson extrapolation). The Wald statistic Z = Δcontrast/SE(Δcontrast) retained all six pairwise sampling covariances among the four correlations. Benjamini–Hochberg correction was applied across the six external traits separately. The component-to-input contrast ratio and its 95% confidence interval were estimated by the same delta method and interpreted only when the 95% confidence interval for the parent SCZ–BIP contrast excluded zero.

#### Supplementary Results

##### Benchmark of ZCA against mtCOJO

Bidirectional mtCOJO conditioning derived SCZ|BIP and BIP|SCZ from the same parent GWASs. Each ZCA component was highly correlated with its mtCOJO counterpart (SCZ_~BIP_ with SCZ|BIP: rg = 0.994, SE = 0.044; BIP_~SCZ_ with BIP|SCZ: rg = 0.996, SE = 0.049). The ZCA components were mutually uncorrelated (rg = −0.025, SE = 0.029), whereas the mtCOJO components retained a modest negative correlation (rg = −0.174, SE = 0.032, P = 5.4 × 10⁻⁸), reflecting the different mathematical targets of whitening and conditional analysis (Supplementary Figure 1). The following result section details the output obtained from the mtCOJO derived component.

##### Genomic risk loci and novelty

FUMA identified 66 loci for SCZ|BIP, 30 for BIP|SCZ and 197 for Psy_shared_. Merging intervals produced 248 consensus loci: 206 were component-specific, 40 were shared by exactly two profiles and two by all three. Six consensus loci contained both conditional components, comprising four shared only by SCZ|BIP and BIP|SCZ and two shared by all three profiles (Supplementary Table 9; Supplementary Figure 3). Ten SCZ|BIP, seven BIP|SCZ and 64 Psy_shared_ loci were not genome-wide significant in either input GWAS. After comparison with the published PGC and Genomic SEM locus sets, one SCZ|BIP locus, two BIP|SCZ loci and 14 Psy_shared_ loci remained absent from every comparison set (Supplementary Table 10).

##### Biological pathways and synaptic enrichment

MAGMA identified 31 FDR-significant gene sets for SCZ|BIP, none for BIP|SCZ and 88 for Psy_shared_ (Supplementary Table 11; Supplementary Figure 4). GABAergic neuron differentiation was significant for SCZ|BIP (P_FDR_ = 0.0195) and Psy_shared_ (P_FDR_ = 0.00171), linking these profiles to neurodevelopmental differentiation. By contrast, GABAergic synapse was significant only for Psy_shared_ (P_FDR_ = 0.00202), consistent with synaptic organisation and signalling rather than neuronal differentiation. SCZ|BIP also showed FDR-significant enrichment for subpallium development, postsynapse and L-type voltage-gated calcium-channel complex; BIP|SCZ showed only nominal pathway signals.

SynGO identified 29 FDR-significant terms for SCZ|BIP, 18 for _BIP|SCZ_ and 29 for Psy_shared_, with 16 terms shared by the two conditional components (Supplementary Table 12). Genesets significant only for SCZ|BIP included postsynaptic-density membrane, postsynaptic-density intracellular component, presynaptic active zone and structural modification of the synapse. The two terms significant only for BIP|SCZ were integral component of the postsynaptic membrane and integral component of the presynaptic membrane. These patterns distinguish synaptic structural organisation from membrane-associated signalling while retaining broad overlap across the profiles.

##### Developmental expression and external genetic correlations:

BrainSpan gene-property analysis showed enrichment during late infancy and young adulthood for both conditional components (SCZ|BIP, P_FDR_ = 0.00410 at each period; BIP|SCZ, P_FDR_ = 0.0222 at each period). Psy_shared_ additionally showed early-mid-prenatal and late-mid-prenatal enrichment (PFDR = 0.00135 and 0.00142), together with late-infancy and young-adulthood enrichment (PFDR = 1.80 × 10⁻⁵ and 0.00135; Supplementary Table 13).

The conditional profiles remained closely aligned with their respective parent disorders (SCZ|BIP–SCZ, rg = 0.920; BIP|SCZ–BIP, rg = 0.877). Against FinnGen endpoints, SCZ|BIP correlated strongly with schizophrenia (rg = 0.910), other psychotic disorders (rg = 0.739) and schizoaffective disorder (rg = 0.660), whereas BIP|SCZ correlated most strongly with bipolar disorder (rg = 0.776). Its correlation with FinnGen schizophrenia was small and did not reach significance (rg = 0.102, P = 0.061). Psy_shared_ showed broad positive correlations and the strongest correlation with the published schizophrenia–bipolar factor (rg = 0.973; Supplementary Table 14; Supplementary Figure 5a,b).

Across intermediate phenotypes, SCZ|BIP showed negative correlations with cognitive task performance (rg = −0.277), educational attainment (rg = −0.037), metabolic syndrome (rg = −0.101), CRP (rg = −0.078) and neutrophil percentage (rg = −0.061), but a positive correlation with risk taking (rg = 0.135). BIP|SCZ showed positive correlations with cognitive task performance (rg = 0.077), educational attainment (rg = 0.132) and risk taking (rg = 0.269); its positive cardiometabolic and inflammatory estimates were not individually significant. Covariance-aware Wald tests showed that the conditional-component contrasts were significantly wider than the input SCZ–BIP contrasts for all six traits after FDR correction (Supplementary Tables 15 and 16; Supplementary Figure 5c,d).

**Supplementary Figure 1 | Genetic correlations across the ZCA and mtCOJO profiles.** Pairwise LDSC genetic correlations (rg) among the SCZ and BIP GWASs, ZCA-derived SCZ_~BIP_ and BIP_~SCZ_, mtCOJO-derived SCZ|BIP and BIP|SCZ, and Psy_shared_. The ZCA components were highly correlated with their mtCOJO counterparts (rg = 0.994 and 0.996) but differed in their inter-component correlation (ZCA: rg = −0.025; mtCOJO: rg = −0.174).

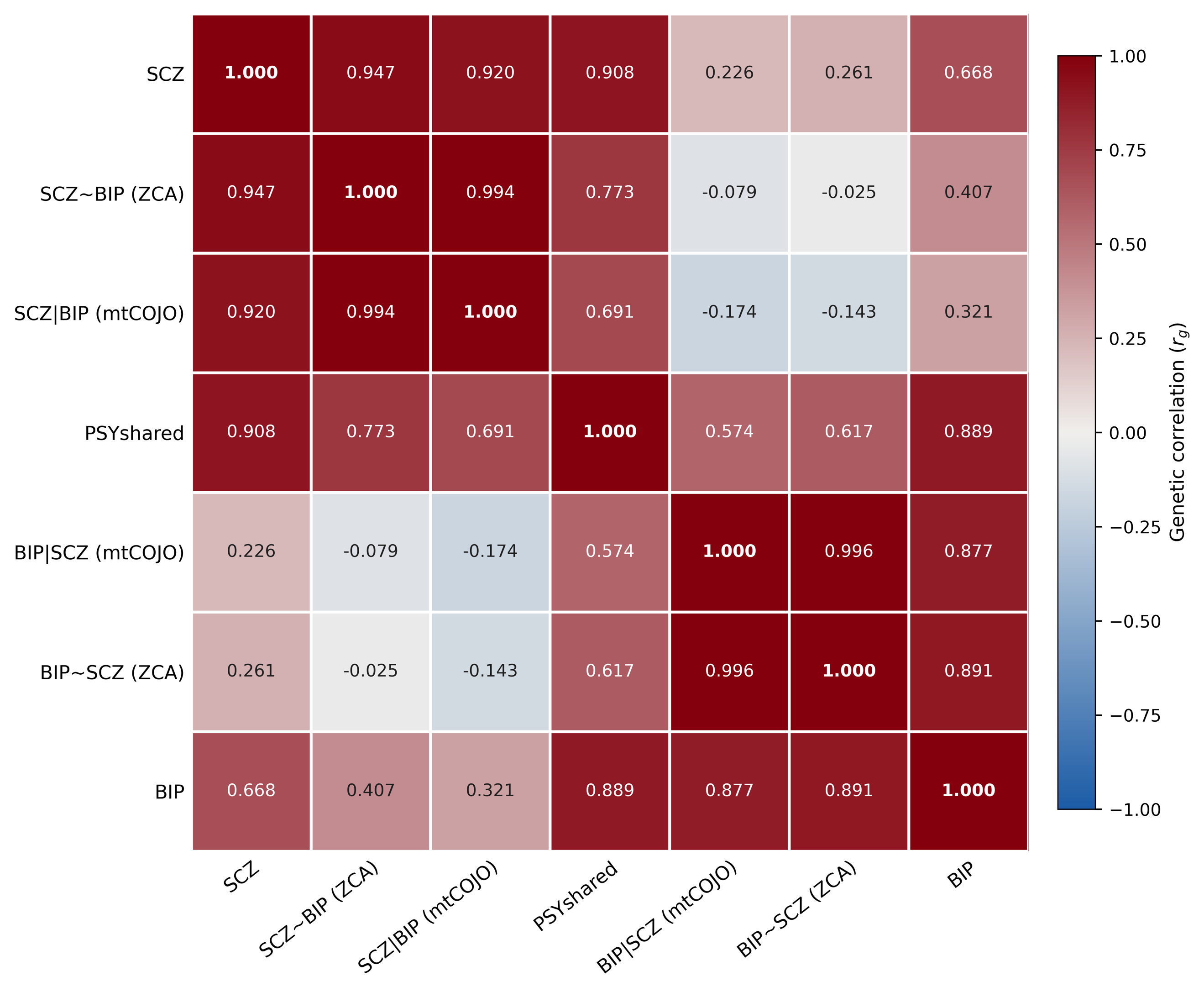

**Supplementary Figure 2 | Study design for the mtCOJO conditional analysis of schizophrenia and bipolar-disorder GWASs and downstream characterisation.** Summary-statistic GWASs of SCZ and BIP were conditioned bidirectionally using mtCOJO to derive SCZ|BIP and BIP|SCZ, while Genomic SEM was used to derive Psy_shared_. Unlike ZCA whitening, mtCOJO does not impose orthogonality. The components were characterised by genomic risk-locus and candidate-gene mapping, novelty assessment, MAGMA pathway and BrainSpan enrichment, SynGO analysis, drug enrichment and druggability annotation, and LDSC genetic correlations with psychiatric, cognitive, cardiometabolic and inflammatory phenotypes. Covariance-aware pairwise tests and Wald comparisons evaluated differences between conditional-component and input-GWAS profiles.

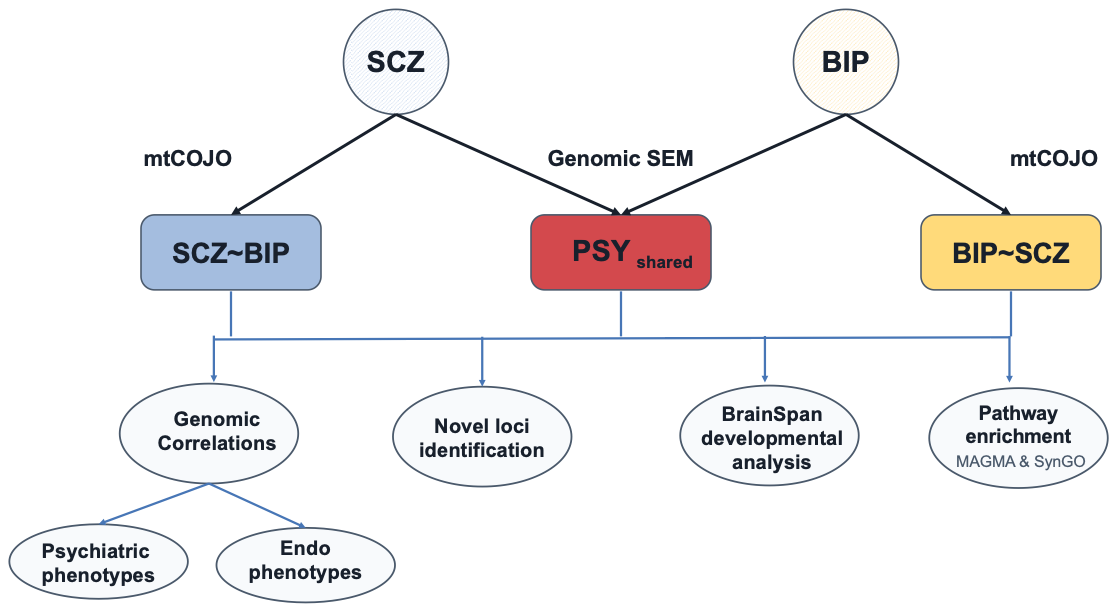

**Supplementary Figure 3a | Manhattan plots for mtCOJO derived component and PSY_shared_.** Manhattan plots show −log10(P) for SCZ|BIP, BIP|SCZ and Psy_shared_. The dashed horizontal line marks genome-wide significance at P = 5 × 10⁻⁸.

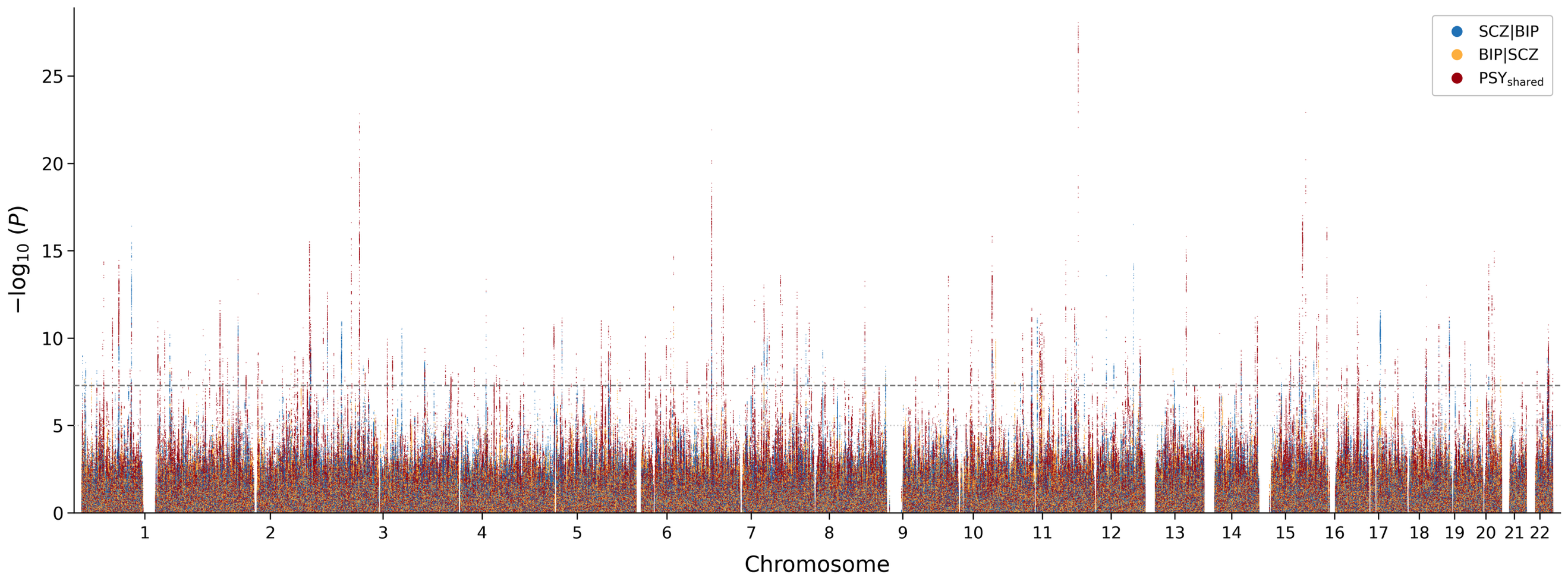

**Supplementary Figure 3b | Consensus genomic risk-locus overlap between mtCOJO derived component and PSY_shared_.** The 66 SCZ|BIP, 30 BIP|SCZ and 197 Psy_shared_ input loci merged at at least 1-bp overlap into 248 consensus loci. Because merging can consolidate multiple input loci, the displayed consensus memberships are 65, 30 and 197, respectively. Of the 248 consensus loci, 206 were component-specific, 40 were shared by exactly two profiles and two by all three. Six loci contained both conditional components: four were shared only by SCZ|BIP and BIP|SCZ, and two by all three profiles. Circle areas are not proportional to counts.

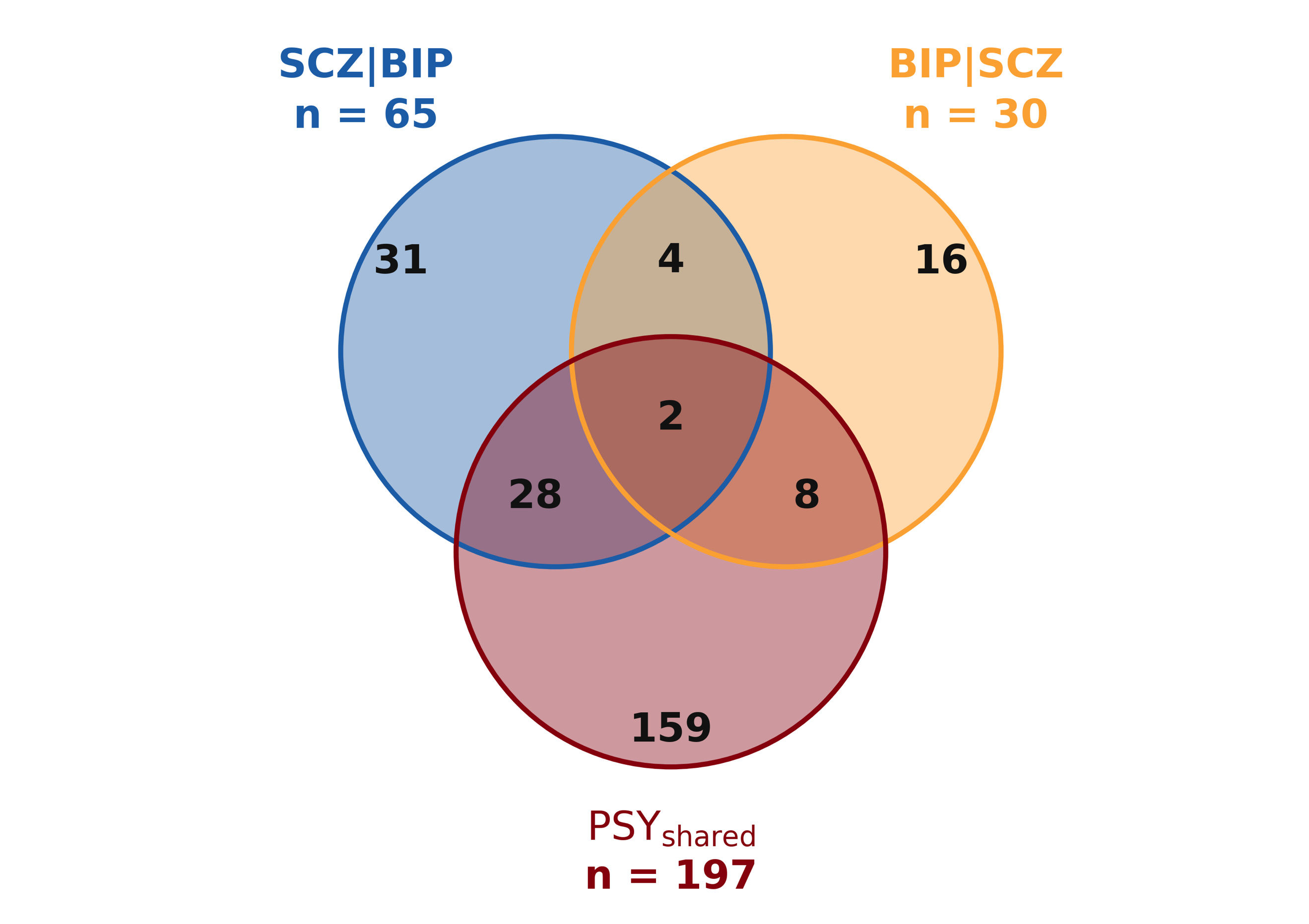

**Supplementary Figure 4 | Biological pathway enrichment across the mtCOJO-component and PSY_shared_ profiles.** MAGMA competitive gene-set enrichment is summarised for SCZ|BIP, BIP|SCZ and Psy_shared_. SCZ|BIP showed FDR-significant neurodevelopmental enrichment including GABAergic neuron differentiation, whereas the GABAergic synapse set was significant only for Psy_shared_ and represents synaptic organisation/signalling rather than neuronal differentiation. BIP|SCZ had no FDR-significant MAGMA gene set; its connections therefore denote nominal signals. Solid lines indicate major enriched groups, dashed lines additional groups with fewer enrichments and thin dotted lines nominal groups that did not reach FDR significance.

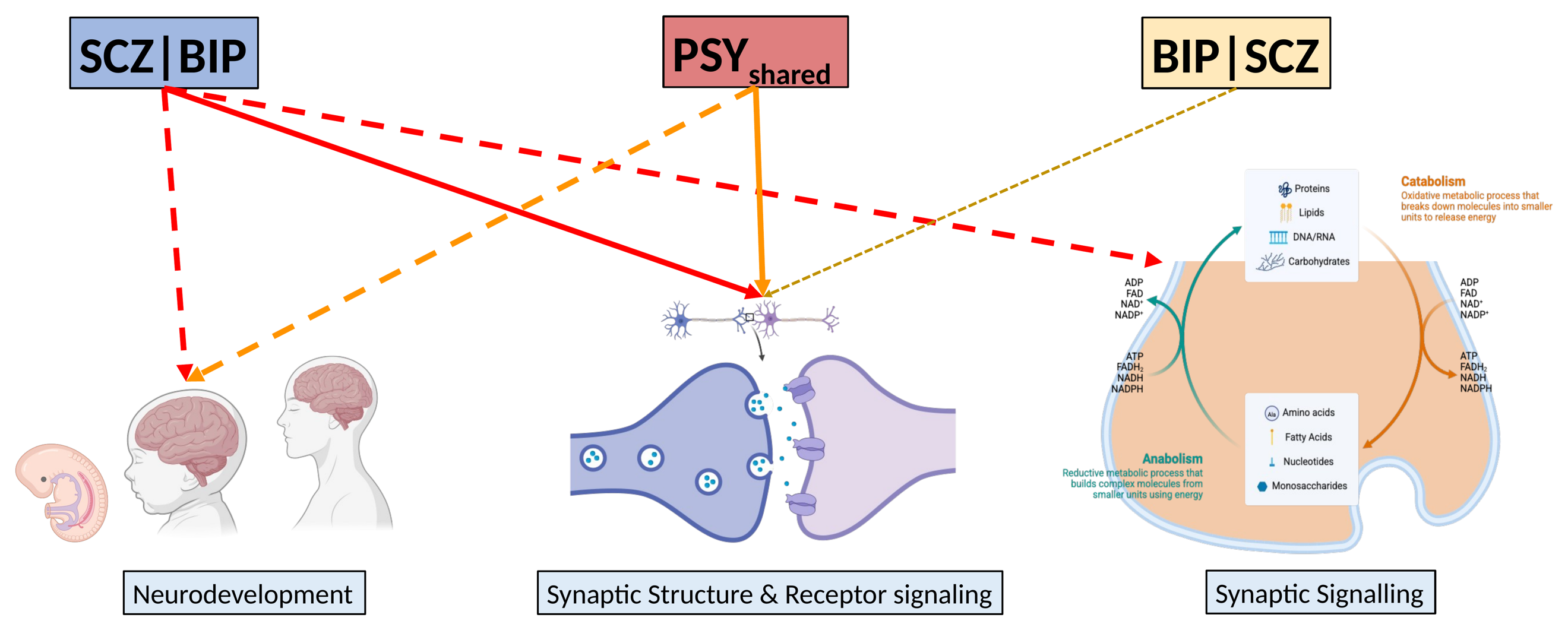

**Supplementary Figure 5a | Pairwise genetic correlations between SCZ, BIP and the mtCOJO derived components.** Pairwise rg values are shown among SCZ, BIP, SCZ|BIP, BIP|SCZ and Psy_shared_. SCZ|BIP remained aligned with SCZ (rg = 0.92), BIP|SCZ with BIP (rg = 0.88), and the conditional components retained a modest negative correlation (rg = −0.17). Both correlated positively with Psy_shared_ (SCZ|BIP, rg = 0.69; BIP|SCZ, rg = 0.57).

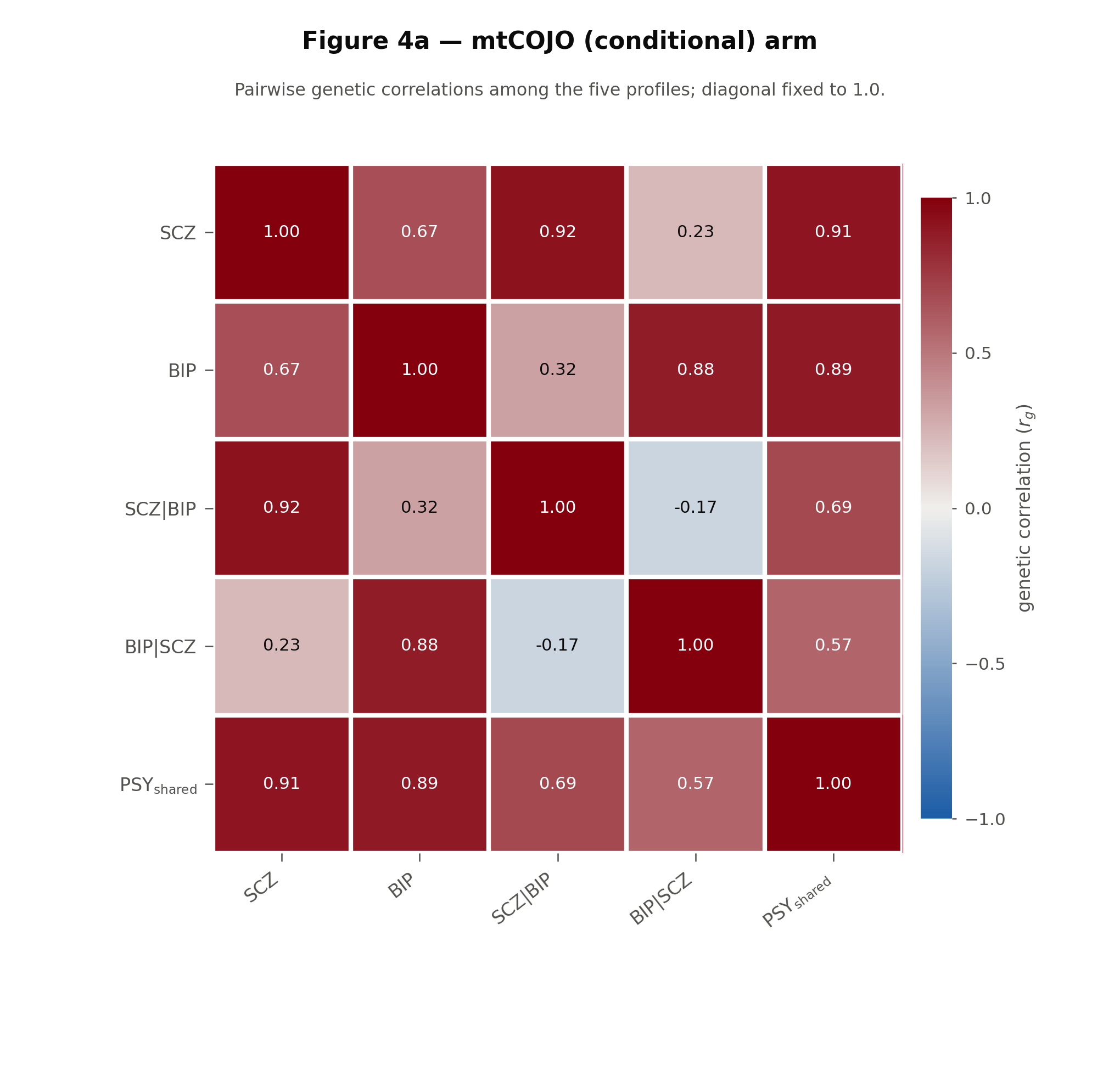

**Supplementary Figure 5b | Genetic correlations with external psychiatric endpoints.** Correlations are shown with FinnGen schizophrenia, schizoaffective disorder, bipolar disorders and other psychotic disorders, together with the schizophrenia–bipolar factor from the 14-disorder Genomic SEM model. SCZ|BIP aligned most strongly with schizophrenia-spectrum endpoints, BIP|SCZ with bipolar disorder, and Psy_shared_ broadly across endpoints. Error bars denote ±1 SE; hatched bars are input GWASs and solid bars are derived profiles.

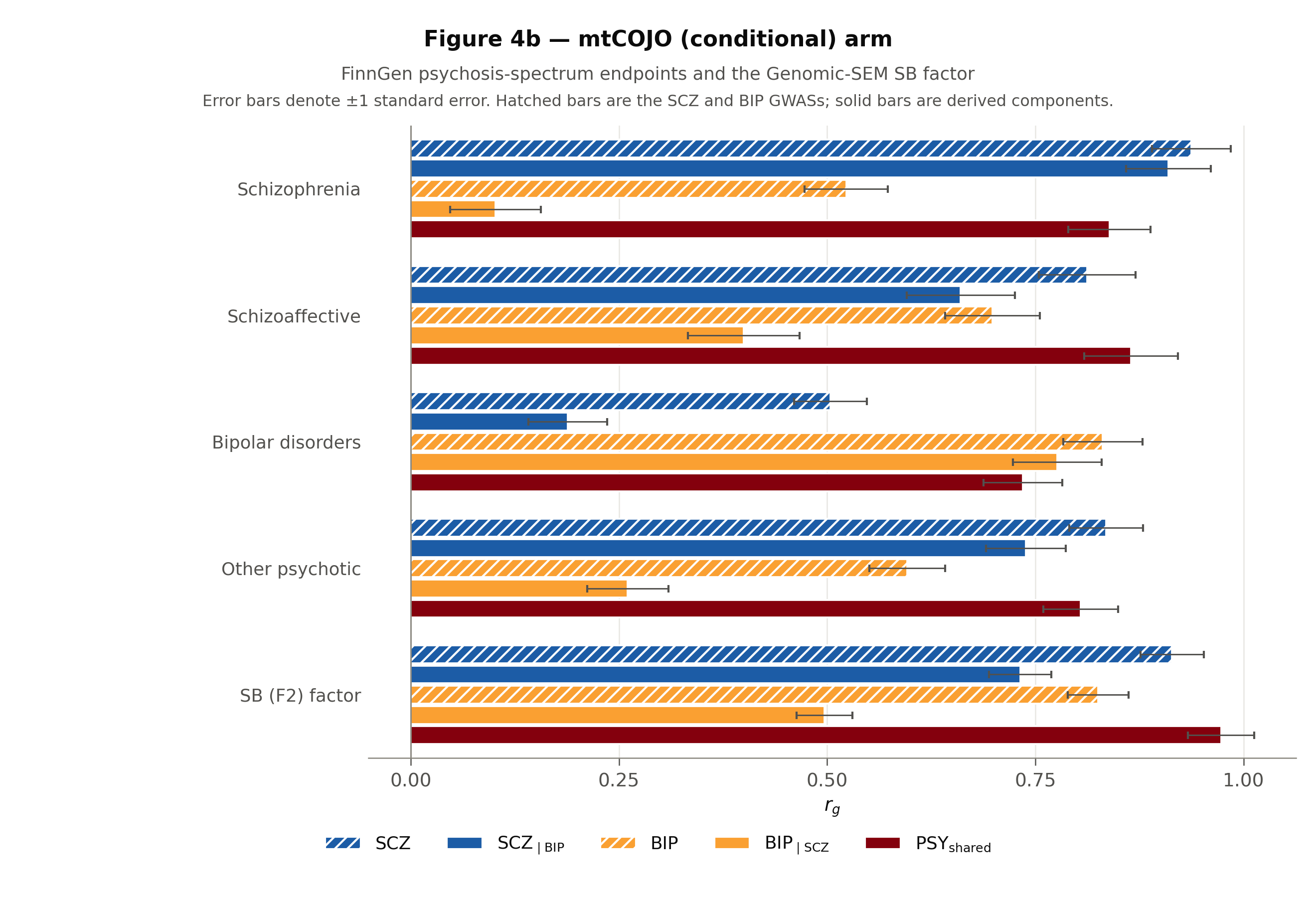

**Supplementary Figure 5c | Genetic correlations with cognitive task performance, educational attainment and risk taking.** SCZ|BIP showed negative cognitive-task and educational-attainment estimates, whereas BIP|SCZ showed positive estimates for both. Psy_shared_ showed a negative cognitive-task estimate and positive educational-attainment and risk-taking estimates. All three derived profiles correlated positively with risk taking. Error bars denote ±1 SE; hatched bars are input GWASs and solid bars are derived profiles.

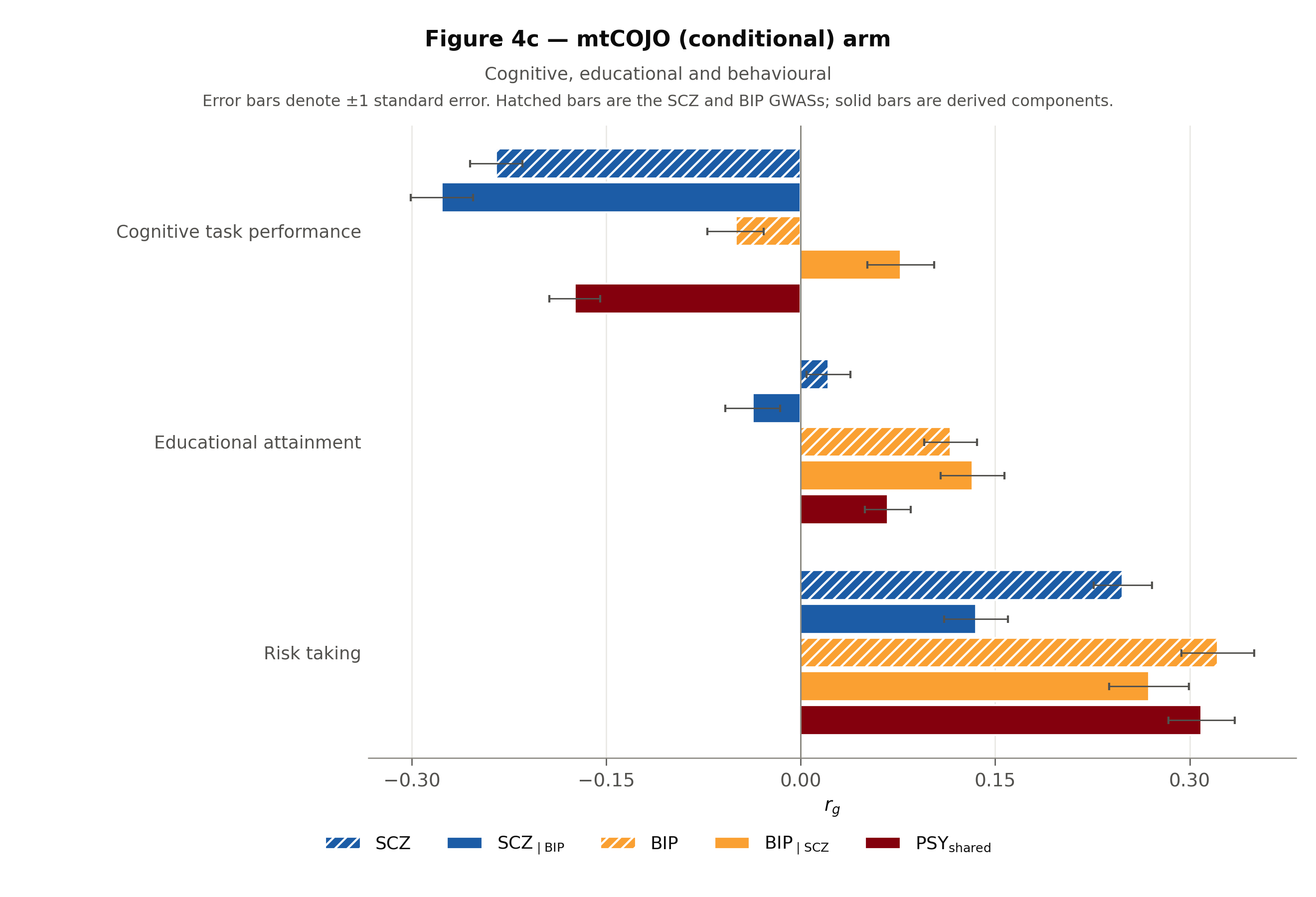

**Supplementary Figure 5d | Genetic correlations with cardiometabolic and inflammatory phenotypes.** SCZ|BIP was significantly negatively correlated with metabolic syndrome, CRP and neutrophil percentage. BIP|SCZ estimates were positive but did not individually reach significance; Psy_shared_ estimates lay between the conditional-component estimates. Error bars denote ±1 SE; hatched bars are input GWASs and solid bars are derived profiles.

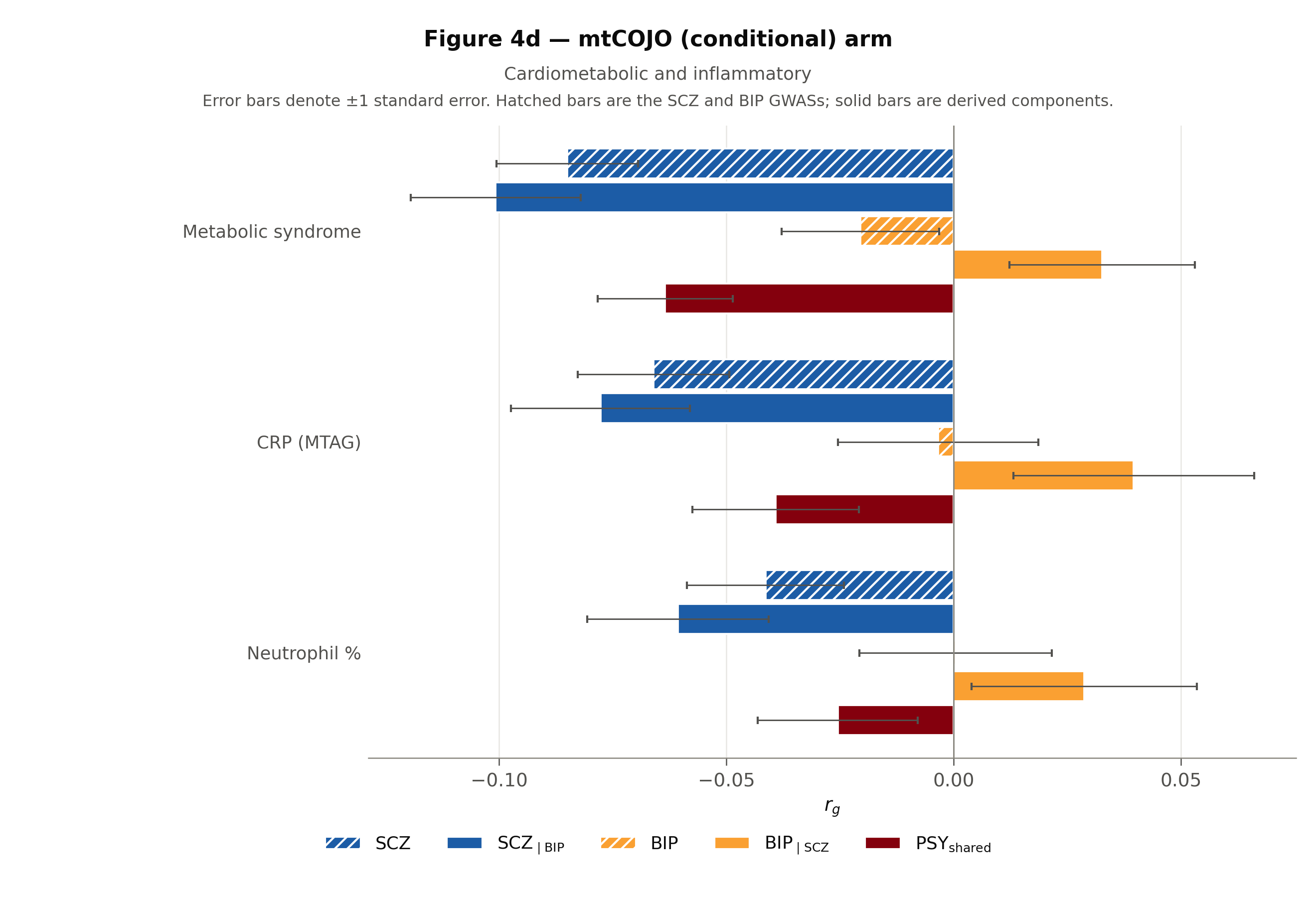
